# Does handgrip strength improve cardiometabolic risk stratification beyond detailed body composition? NHANES 2011–2014 with temporal validation

**DOI:** 10.64898/2026.10.09.26365176

**Authors:** Xiaonan Zhan, Majun Shen, Yichi Zhang, Xingwei Mao

**Affiliations:** Monash University, School of Information Technology; China Jiliang University, Hangzhou, China; National University of Singapore, Singapore

**Author notes:** **Corresponding author:** Xiaonan Zhan.

**Keywords:** Handgrip strength, Body composition, Visceral adipose tissue, Cardiometabolic risk, Incremental value, Temporal validation, Machine learning

## Abstract

**Background:** Handgrip strength is associated with cardiometabolic abnormalities, but whether it adds information once detailed adiposity and muscle mass are known is unclear.

**Methods:** We studied adults aged 40–59 years from NHANES 2011–2012 (development, n=508) and 2013–2014 (temporal validation, n=616). The outcome was at least two of four non-adiposity abnormalities (elevated blood pressure, fasting glucose or triglycerides, or low HDL cholesterol); waist circumference was not part of the outcome. A prespecified feature ladder added DXA body fat percentage and visceral adipose tissue to age, sex, BMI and waist circumference (M1), then appendicular skeletal muscle index (M2), then absolute (M3A, primary) or weight-normalised (M3B) grip strength. Logistic regression was primary; elastic-net and XGBoost were supportive. Development performance used nested cross-validation; frozen models were scored once in the validation cohort. Paired increments used 2000 stratified bootstrap resamples.

**Results:** The outcome occurred in 249 (49.0%) and 277 (45.0%) participants. Adding adiposity increased the logistic regression AUC by 0.0209 (95% CI 0.0024–0.0402) in development and 0.0209 (0.0032–0.0375) in temporal validation. Adding absolute grip strength to M2 changed the AUC by −0.0046 (−0.0107 to 0.0013) and 0.0045 (−0.0009 to 0.0098), respectively. All 12 grip-strength AUC increments across algorithms, definitions and cohorts had intervals that included zero. Calibration was unchanged and the incremental net benefit was not consistently positive. Model attributions to grip strength varied by algorithm.

**Conclusions:** In middle-aged adults, detailed adiposity, but not grip strength, added reproducible information for concurrent cardiometabolic risk stratification beyond conventional anthropometry and muscle mass.

**Highlights:**

- DXA adiposity added about 0.02 AUC in both development and temporal validation
- Grip strength added no stable increment beyond adiposity and muscle mass
- All 12 grip-strength AUC increments had 95% CIs that included zero
- Grip strength received model attributions that did not improve performance
- Frozen models were scored once in a later, temporally separated NHANES cycle

## 1. Introduction

Cardiometabolic abnormalities, including elevated blood pressure, dysglycaemia, hypertriglyceridaemia and low high-density lipoprotein (HDL) cholesterol, tend to occur together and underlie much of the burden of cardiovascular disease and type 2 diabetes[1]. Body mass index (BMI) and waist circumference are the usual clinical indicators of excess adiposity, but neither separates fat from lean mass or visceral from subcutaneous fat. Visceral adipose tissue (VAT) is more strongly related to metabolic risk factors than subcutaneous fat after accounting for BMI and waist circumference [2], and it is associated with incident prediabetes and diabetes in adults with obesity [3]. Detailed body composition may therefore carry information that anthropometry does not.

Skeletal muscle is the other major body compartment. Low muscle mass has been associated with insulin resistance and dysglycaemia [4], although in older men sarcopenic obesity did not carry a greater risk of the metabolic syndrome than obesity alone [5]. Handgrip strength, a simple and inexpensive measure of muscle function, is inversely associated with cardiovascular events and mortality in large cohorts [6,7]. Adding grip strength to established risk scores has produced small improvements in the prediction of cardiovascular disease and type 2 diabetes [7–9], and cross-sectional studies have reported that low relative grip strength is associated with the metabolic syndrome [10,11].

An association, or good discrimination by a marker on its own, does not show that the marker improves an existing model [12]. For risk stratification, the relevant question is whether grip strength adds information once adiposity and muscle quantity are already known. Few studies have examined this with a prespecified sequence of models, paired comparisons in the same participants and validation in a later sample. Machine-learning analyses of such questions are also prone to optimistic bias when outcome components enter the predictor set, when preprocessing or tuning uses validation data, or when models are chosen after test results have been seen [13,14].

We therefore analysed two consecutive cycles of the National Health and Nutrition Examination Survey (NHANES) under leakage controls. Using a prespecified feature ladder, we asked whether dual-energy X-ray absorptiometry (DXA) adiposity measures improve stratification beyond age, sex, BMI and waist circumference; whether appendicular muscle mass adds further; and whether absolute or weight-normalised grip strength adds beyond all of these. The primary comparison was the addition of absolute grip strength to a logistic regression model that already contained anthropometry, DXA adiposity and muscle mass, evaluated with frozen models in a temporally separated validation cohort. The adiposity step is examined in more detail, with VAT and body fat percentage assessed separately, in a companion analysis of NHANES 2015–2018 (manuscript in preparation); the present study focuses on muscle function.

## 2. Methods

### 2.1. Study design and data source

NHANES is a continuous cross-sectional survey of the civilian, non-institutionalised US population. It uses a complex, multistage probability design and combines household interviews with standardised examinations and laboratory tests in mobile examination centres [15]. We used the 2011–2012 cycle for model development and the 2013–2014 cycle as a temporally separated validation cohort. Because predictors and outcome were measured at the same visit, the models address concurrent risk stratification (phenotype classification) rather than prediction of future events. The analysis plan, covering the outcome, predictor whitelist, feature ladder, algorithms, cross-validation scheme, comparisons, metrics and bootstrap settings, was frozen before model training (26 August 2026), and its machine-readable specification was protected with a SHA-256 checksum; the study was not registered. Reporting follows the TRIPOD+AI statement [16] (Supplementary Table S6).

### 2.2. Participants

In each cycle we identified non-pregnant participants aged 20–59 years with complete anthropometry, a valid whole-body DXA examination, valid grip strength, blood pressure and HDL cholesterol measurements, and fasting-subsample laboratory data (fasting glucose and triglycerides after an 8–24 h fast). Pregnancy was identified from the examination pregnancy status or a positive urine pregnancy test. The prespecified primary analysis was restricted to participants aged 40–59 years; the upper limit reflects the age range of the NHANES whole-body DXA examination (Fig. 1A). Participants were not excluded because of missing predictor values.

**Fig. 1.**
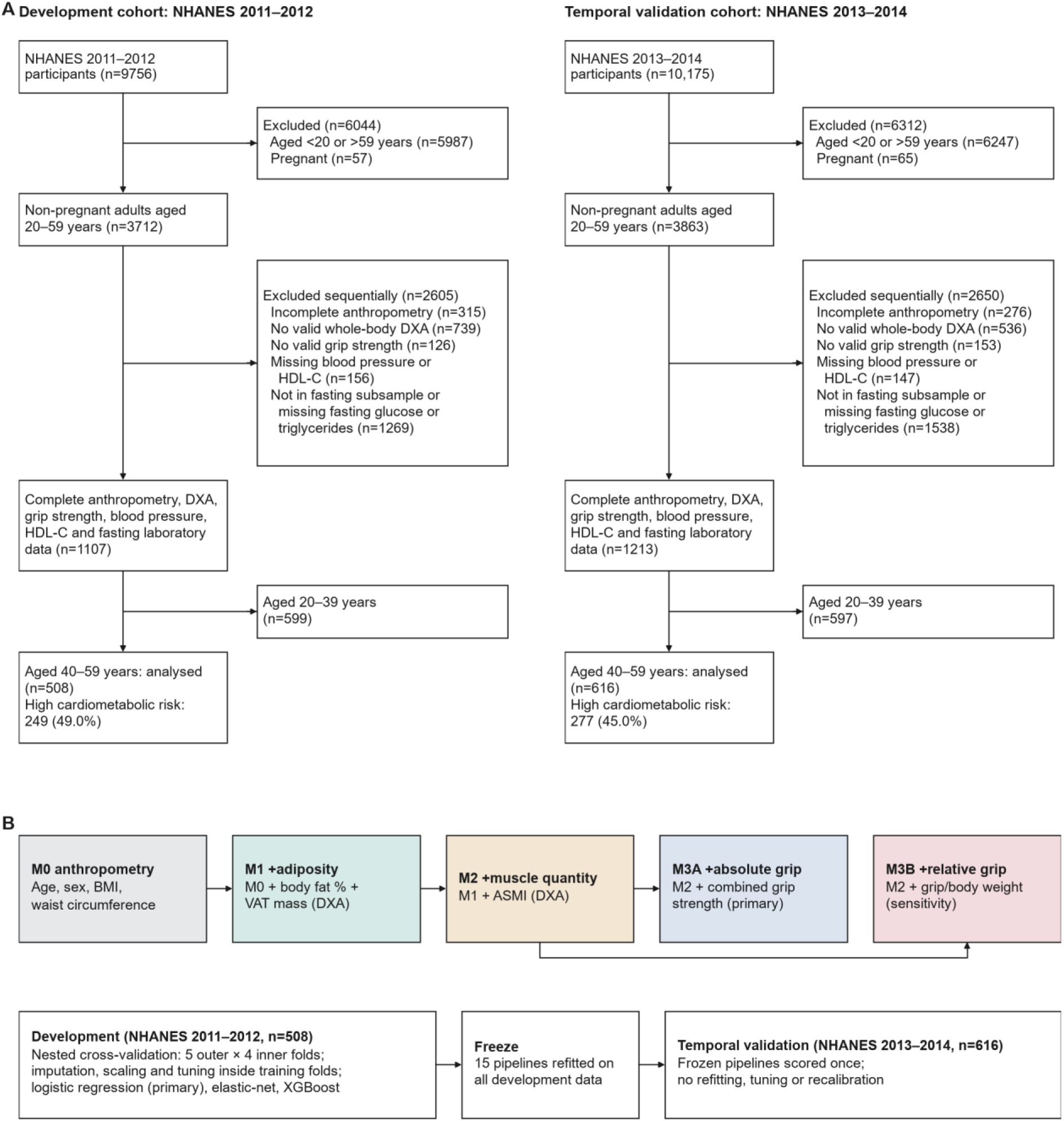
Participant flow and study design. (A) Participant flow in the development (NHANES 2011–2012) and temporal validation (NHANES 2013–2014) cohorts; exclusions after the second box were applied sequentially in the order listed. All participants with complete fasting-subsample data had fasted 8–24 h, so the fasting criterion excluded no one. (B) Prespecified feature ladder and development–validation workflow. Shading in panel B only distinguishes the steps of the ladder and has no statistical meaning. ASMI=appendicular skeletal muscle index. DXA=dual-energy X-ray absorptiometry. VAT=visceral adipose tissue.

### 2.3. Outcome

The primary outcome, high cardiometabolic risk, required at least two of four non-adiposity abnormalities adapted from harmonised metabolic syndrome criteria [1]: systolic blood pressure ≥130 mmHg, diastolic blood pressure ≥85 mmHg or current antihypertensive medication; fasting glucose ≥100 mg/dL, diagnosed diabetes or use of insulin or oral glucose-lowering drugs; triglycerides ≥150 mg/dL; and HDL cholesterol <40 mg/dL in men or <50 mg/dL in women. Blood pressure was the mean of the second to fourth readings when available. Waist circumference, a component of the metabolic syndrome, was deliberately excluded from the outcome because it was also a predictor, which would otherwise have created definitional overlap between predictors and outcome.

### 2.4. Predictors and feature ladder

Predictors entered the models only through a fixed whitelist. M0 contained age, sex, BMI and waist circumference. M1 added DXA whole-body fat percentage and VAT mass [17,18]. M2 added the appendicular skeletal muscle index (ASMI; arm and leg lean mass divided by height squared) [19]. M3A, the primary model, added absolute grip strength, defined by NHANES as the sum of the largest reading from each hand (combined grip strength) [20]; M3B, a sensitivity model, instead added grip strength divided by body weight. Absolute and relative grip strength never entered the same model. Blood pressure, glucose, triglyceride and HDL values, diagnoses, medication use, outcome components, survey design variables and identifiers were excluded from all predictor sets. Variable definitions are given in Supplementary Methods S1.

### 2.5. Model development

Three model families were fitted for every feature set: logistic regression with a fixed, weak L2 penalty (C=1 on standardised predictors, without tuning), prespecified as the primary model; elastic-net logistic regression [21]; and gradient-boosted trees (XGBoost) with strong regularisation [22]. Preprocessing was embedded in each pipeline and learned only within training folds: continuous predictors were median-imputed and standardised, and sex was mode-imputed and one-hot encoded. No class-imbalance methods were used because the outcome prevalence was close to 50%. Models output predicted probabilities.

Development performance was estimated by nested cross-validation with five stratified outer folds and four stratified inner folds within each outer training fold [13]. Hyperparameters of elastic-net (30 random candidates) and XGBoost (40 random candidates) were tuned in the inner folds with the AUC as the only refit criterion, and every participant received one out-of-fold (OOF) prediction from each model. Search spaces and selected values are listed in Supplementary Table S5. After the development analyses were complete, each of the 15 pipelines (five feature sets by three algorithms) was refitted on all 508 development participants, with hyperparameters selected by four-fold cross-validation, and frozen.

### 2.6. Temporal validation and data-access history

The frozen pipelines were applied once to the 2013–2014 cohort, without refitting, tuning, threshold selection or recalibration. The validation files had been assembled together with the development data, and before any model was fitted their outcome prevalence, predictor distributions and missingness were summarised as part of data-quality checks; the files were then locked with file-system access controls until the models and evaluation code had been frozen. During the single scoring run, two attempts stopped because of numerical convergence failures in the calibration diagnostics, before any performance estimate had been produced; the calibration routine was made numerically robust without changing models, predictions or thresholds, and the run was then completed (three reads of the validation matrix in total). After validation, the locked files were opened once more, with the investigators’ authorisation, to extract descriptive characteristics, participant-flow counts and survey design variables for Table 1 and the weighted sensitivity analysis; no predictions were recomputed and no model was changed. We therefore describe 2013–2014 as a temporally separated validation cohort rather than an untouched external test set (Supplementary Methods S2).

**Table 1.** Characteristics of the development and temporal validation cohorts.

| Characteristic | Development<br>NHANES 2011–2012<br>(n=508) | Temporal validation<br>NHANES 2013–2014<br>(n=616) | SMD |
| --- | --- | --- | --- |
| Age, years | 49.3 (5.8) | 49.0 (5.7) | 0.06 |
| Female sex | 246 (48.4) | 312 (50.6) | −0.04 |
| **Race and ethnicity** |  |  |  |
| Mexican American | 52 (10.2) | 86 (14.0) | −0.11 |
| Other Hispanic | 49 (9.6) | 63 (10.2) | −0.02 |
| Non-Hispanic White | 197 (38.8) | 260 (42.2) | −0.07 |
| Non-Hispanic Black | 132 (26.0) | 108 (17.5) | 0.21 |
| Non-Hispanic Asian | 60 (11.8) | 84 (13.6) | −0.05 |
| Other or multiracial | 18 (3.5) | 15 (2.4) | 0.07 |
| Family income-to-poverty ratio | 2.76 (1.69) | 2.82 (1.69) | −0.04 |
| **Body size and composition** |  |  |  |
| BMI, kg/m<sup>2</sup> | 29.5 (6.6) | 29.2 (6.5) | 0.05 |
| Waist circumference, cm | 100.4 (15.7) | 99.2 (14.6) | 0.08 |
| Body fat, % | 34.0 (8.4) | 34.1 (8.2) | 0.00 |
| VAT mass, g | 596 (282) | 589 (282) | 0.02 |
| ASMI, kg/m<sup>2</sup> | 7.97 (1.61) | 7.82 (1.61) | 0.09 |
| **Muscle function** |  |  |  |
| Combined grip strength, kg | 73.3 (19.8) | 73.3 (20.3) | 0.00 |
| Grip strength/body weight, kg/kg | 0.90 (0.24) | 0.92 (0.25) | −0.06 |
| **Outcome components** |  |  |  |
| Elevated blood pressure | 240 (47.2) | 253 (41.1) | 0.12 |
| Elevated fasting glucose | 275 (54.1) | 310 (50.3) | 0.08 |
| Elevated triglycerides | 159 (31.3) | 158 (25.6) | 0.13 |
| Low HDL cholesterol | 135 (26.6) | 167 (27.1) | −0.01 |
| High cardiometabolic risk (≥2 components) | 249 (49.0) | 277 (45.0) | 0.08 |
| **Clinical measurements** |  |  |  |
| Systolic blood pressure, mmHg | 122.7 (15.6) | 121.1 (16.4) | 0.10 |
| Diastolic blood pressure, mmHg | 74.2 (10.5) | 71.4 (10.5) | 0.27 |
| Fasting glucose, mg/dL, median (IQR) | 100 (93–111) | 99 (94–109) | 0.02 |
| Triglycerides, mg/dL, median (IQR) | 111 (77–167) | 101 (68–150) | 0.16 |
| HDL cholesterol, mg/dL | 53.3 (16.2) | 53.8 (15.9) | −0.03 |
| Antihypertensive medication | 134 (26.4) | 141 (22.9) | 0.08 |
| Insulin or glucose-lowering pills | 62 (12.2) | 57 (9.3) | 0.10 |

Values are unweighted mean (SD), median (interquartile range) or n (%). SMD is the standardised mean difference between cohorts (development minus validation); for medians it was computed from means and SDs. The family income-to-poverty ratio was missing for 33 (6.5%) and 52 (8.4%) participants and VAT mass for 4 (0.8%) and 2 (0.3%); all other variables were complete. Outcome components are defined in Section 2.3. Survey-weighted outcome prevalence was 45.9% (95% CI 40.0–53.3) in development and 43.9% (37.6–51.1) in validation. ASMI=appendicular skeletal muscle index. BMI=body mass index. HDL=high-density lipoprotein. IQR=interquartile range. VAT=visceral adipose tissue.

### 2.7. Performance evaluation

Discrimination was assessed with the area under the receiver operating characteristic curve (AUC; primary metric) and average precision, and overall accuracy with the Brier score and log loss. Calibration was summarised by calibration-in-the-large (CITL; the intercept with the slope fixed at 1), the calibration slope and calibration curves over ten equal-size groups [23]. Clinical utility was assessed with decision curves over the prespecified threshold range 0.05–0.50 in steps of 0.01 [24]. Increments were calculated as the extended model minus the reference model; for the Brier score and log loss they are reported as reductions, so that positive values indicate improvement. Uncertainty was quantified with 2000 stratified, paired, participant-level bootstrap resamples of the fixed predictions (percentile 95% confidence intervals [CIs]), so that both models in a comparison were evaluated on the same resampled participants. A confirmatory claim for grip strength required a positive paired ΔAUC for M3A versus M2, no material worsening of the Brier score or calibration, and assessment of net benefit across the threshold range. The other comparisons were secondary or sensitivity analyses; no adjustment for multiplicity was made and P values were not used for inference.

### 2.8. Model explanation

To describe how the models used grip strength, we computed cross-fitted explanations in the development cohort: each participant’s contributions came from the outer-fold model that had not been trained on that participant. For logistic regression, contributions were the standardised coefficient multiplied by the standardised predictor value on the log-odds scale, and coefficient stability was examined across the five outer folds; for XGBoost we used TreeSHAP [25,26]. The cross-fitted contributions reproduced the OOF predictions (maximum absolute error in predicted probability 1.1×10⁻¹⁶). Attributions describe model behaviour and were not interpreted as incremental value or causal effects.

### 2.9. Descriptive statistics and survey-weighted sensitivity analysis

Cohort characteristics are reported as unweighted mean (SD), median (interquartile range) or n (%), with standardised mean differences (SMDs) [27]. Because NHANES oversamples some population groups, the primary analyses describe model performance in the analysed samples rather than in the US population. In a sensitivity analysis, we re-estimated AUCs, Brier scores and paired increments with the fasting-subsample weights, masked variance strata and pseudo-primary sampling units (PSUs) [28]. CIs were obtained with a Rao–Wu rescaling bootstrap (2000 replicates; in each stratum with n PSUs, n−1 were sampled with replacement) [29] applied to the frozen predictions. As a check, bootstrap standard errors agreed with Taylor linearisation for the weighted outcome prevalence in the development cohort (0.036 vs 0.035).

### 2.10. Software and ethics

Analyses used Python 3.12 with scikit-learn 1.7.2 and XGBoost 3.4.1; figures were produced with matplotlib to Elsevier artwork specifications. Analysis scripts are numbered by step and archived with checksums of inputs and outputs. The NHANES protocols were approved by the National Center for Health Statistics Ethics Review Board (2011–2012: Protocol #2011-17; 2013–2014: continuation of Protocol #2011-17), and all participants provided informed consent [30]. Patients and the public were not involved in the design or conduct of this secondary analysis. This secondary analysis used only de-identified, publicly available NHANES data.

## 3. Results

### 3.1. Participants

Of 9756 participants in NHANES 2011–2012 and 10,175 in NHANES 2013–2014, 508 and 616 aged 40–59 years met the inclusion criteria (Fig. 1A). The outcome was present in 249 (49.0%) and 277 (45.0%) participants, corresponding to survey-weighted prevalences of 45.9% (95% CI 40.0–53.3) and 43.9% (37.6–51.1). The two cohorts were similar in age, sex, body size, body composition and grip strength, with absolute SMDs of 0.09 or less for every predictor (Table 1). The validation cohort had fewer non-Hispanic Black participants (17.5% vs 26.0%), lower diastolic blood pressure and triglyceride levels, and lower prevalences of elevated blood pressure (41.1% vs 47.2%) and elevated triglycerides (25.6% vs 31.3%). VAT mass was missing for four (0.8%) and two (0.3%) participants; all other predictors were complete. Within each cohort, participants with the outcome had higher BMI, waist circumference, VAT mass and ASMI; their absolute grip strength was higher and their relative grip strength lower, consistent with a higher proportion of men (Supplementary Table S1).

### 3.2. Discrimination across the feature ladder

In the development cohort, the logistic regression AUC was 0.748 (95% CI 0.704–0.789) for M0, 0.768 (0.727–0.809) for M1, 0.766 (0.725–0.807) for M2 and 0.761 (0.719–0.802) for M3A (Table 2; Fig. 2A). In temporal validation, the corresponding values were 0.722 (0.681–0.762), 0.743 (0.703–0.781), 0.742 (0.702–0.780) and 0.746 (0.707–0.785) (Fig. 2B). Across all 15 models, AUCs ranged from 0.737 to 0.772 in development and from 0.722 to 0.748 in validation; differences between algorithms were smaller than the difference between M0 and M1 (Supplementary Fig. S2; Supplementary Table S2).

**Fig. 2.**
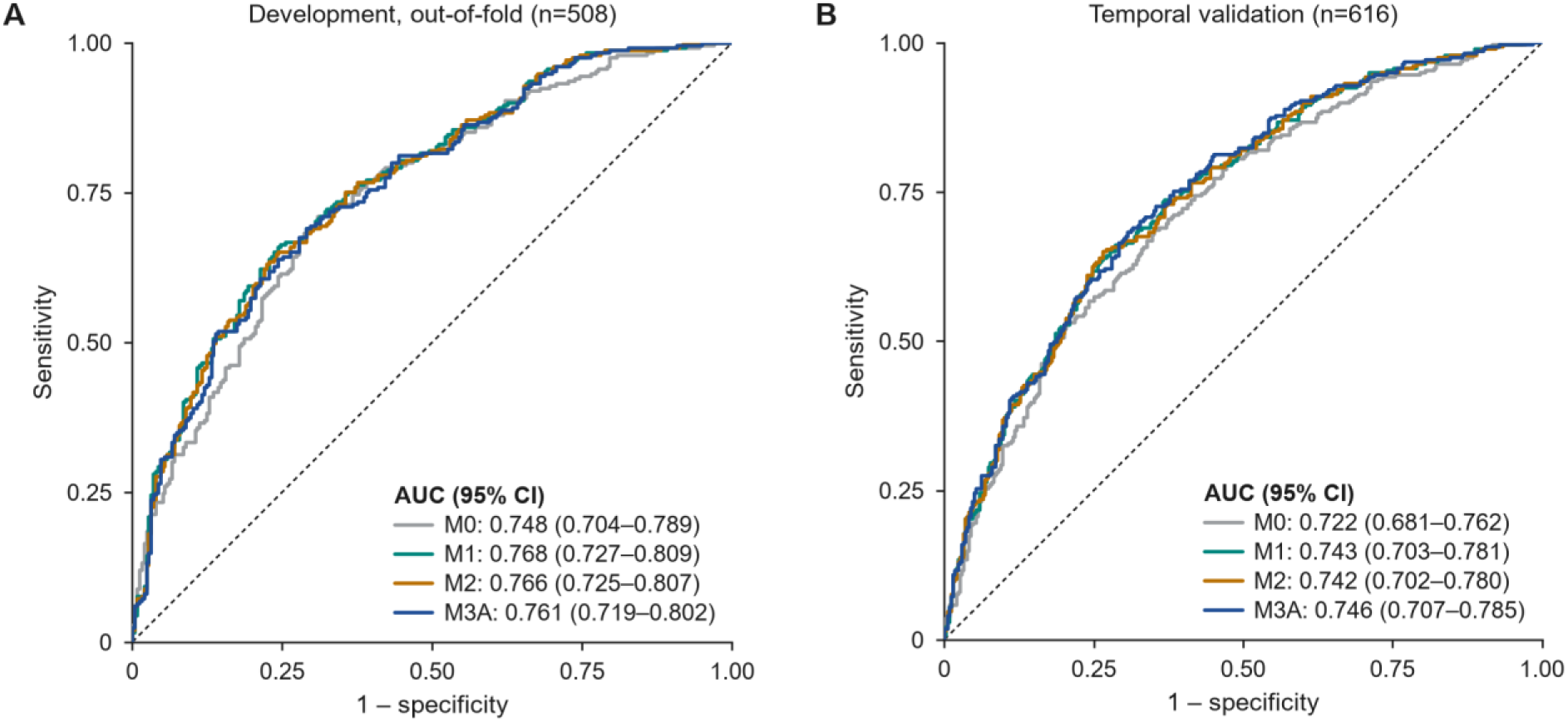
Discrimination of the logistic regression feature ladder. Receiver operating characteristic curves for M0 (age, sex, BMI and waist circumference), M1 (M0 plus body fat percentage and VAT mass), M2 (M1 plus ASMI) and M3A (M2 plus absolute grip strength). (A) Out-of-fold predictions from nested cross-validation in the development cohort (n=508). (B) Frozen models scored once in the temporal validation cohort (n=616). AUCs are empirical; 95% CIs are percentiles of 2000 stratified bootstrap resamples. The dashed line is the line of no discrimination. AUC=area under the receiver operating characteristic curve.

**Table 2.** Performance of the logistic regression feature ladder.

| Model | AUC (95% CI) | Average precision | Brier score | CITL (95% CI) | Calibration slope (95% CI) |
| --- | --- | --- | --- | --- | --- |
| **Development, out-of-fold (n=508; 249 events)** |  |  |  |  |  |
| M0: age, sex, BMI, waist | 0.748 (0.704–0.789) | 0.735 | 0.204 | 0.00 (–0.09 to 0.09) | 0.95 (0.74–1.18) |
| M1: M0 + body fat %, VAT | 0.768 (0.727–0.809) | 0.752 | 0.196 | 0.00 (–0.10 to 0.10) | 0.93 (0.74–1.16) |
| M2: M1 + ASMI | 0.766 (0.725–0.807) | 0.749 | 0.197 | 0.00 (–0.10 to 0.10) | 0.90 (0.72–1.13) |
| M3A: M2 + absolute grip | 0.761 (0.719–0.802) | 0.744 | 0.199 | –0.01 (–0.11 to 0.09) | 0.88 (0.70–1.11) |
| M3B: M2 + relative grip | 0.763 (0.722–0.805) | 0.746 | 0.198 | 0.00 (–0.11 to 0.09) | 0.89 (0.71–1.12) |
| **Temporal validation (n=616; 277 events)** |  |  |  |  |  |
| M0: age, sex, BMI, waist | 0.722 (0.681–0.762) | 0.663 | 0.213 | –0.12 (–0.20 to –0.04) | 0.84 (0.65–1.04) |
| M1: M0 + body fat %, VAT | 0.743 (0.703–0.781) | 0.684 | 0.206 | –0.15 (–0.24 to –0.06) | 0.81 (0.64–1.00) |
| M2: M1 + ASMI | 0.742 (0.702–0.780) | 0.686 | 0.206 | –0.14 (–0.24 to –0.05) | 0.80 (0.64–0.99) |
| M3A: M2 + absolute grip | 0.746 (0.707–0.785) | 0.690 | 0.204 | –0.13 (–0.22 to –0.04) | 0.81 (0.65–1.00) |
| M3B: M2 + relative grip | 0.745 (0.705–0.783) | 0.689 | 0.205 | –0.13 (–0.23 to –0.04) | 0.80 (0.64–1.00) |

Development estimates are from out-of-fold predictions of nested cross-validation; temporal validation estimates are from frozen models scored once, without recalibration. 95% CIs are percentiles of 2000 stratified bootstrap resamples. CITL is the calibration intercept with the slope fixed at 1 (negative values indicate overestimation). AUC=area under the receiver operating characteristic curve. ASMI=appendicular skeletal muscle index. CITL=calibration-in-the-large. VAT=visceral adipose tissue.

### 3.3. Incremental value

Adiposity (M1 vs M0) was the only step with a consistent increment (Table 3; Fig. 3). With logistic regression, ΔAUC was 0.0209 (0.0024–0.0402) in development and 0.0209 (0.0032–0.0375) in validation, and the Brier score improved by 0.0081 (0.0006–0.0155) and 0.0069 (0.0004–0.0131). XGBoost gave similar AUC increments of 0.0246 (0.0007–0.0488) and 0.0243 (0.0045–0.0433), whereas the elastic-net intervals included zero. Adding ASMI (M2 vs M1) did not improve discrimination in any model; with XGBoost in validation, the AUC changed by −0.0112 (−0.0218 to −0.0007).

**Fig. 3.**
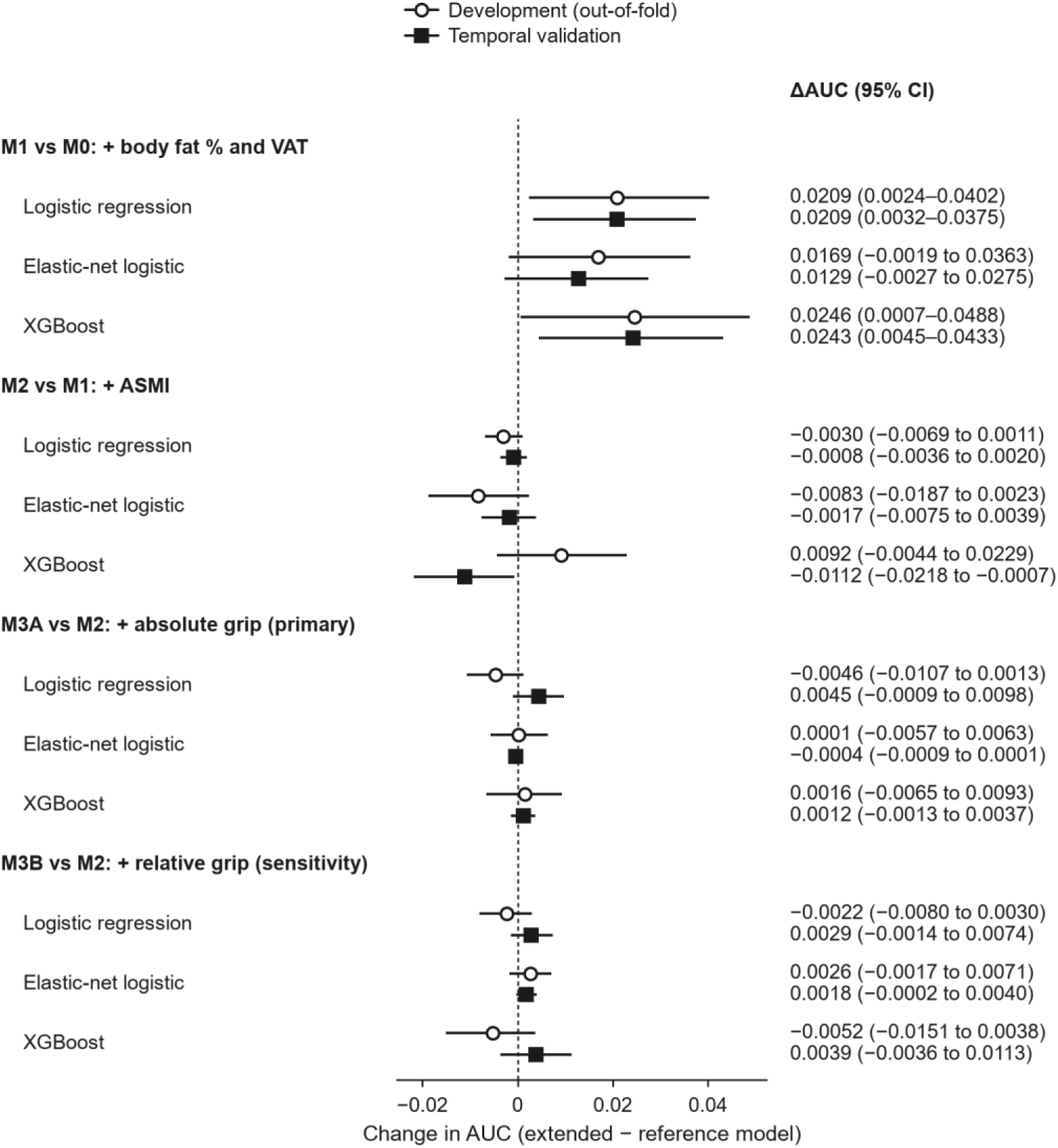
Paired change in AUC for the prespecified comparisons. Each row shows the change in AUC when the extended model is compared with its reference model on the same participants, for three algorithms. Open circles show development out-of-fold estimates (n=508) and filled squares temporal validation estimates (n=616); horizontal lines are 95% CIs from 2000 stratified paired bootstrap resamples. The dashed line marks no change. The primary comparison was M3A vs M2 with logistic regression; the other comparisons are secondary or sensitivity analyses, and no adjustment for multiplicity was made.

**Table 3.** Paired changes in AUC and Brier score for the prespecified comparisons.

| Comparison and algorithm | ΔAUC,<br>development | ΔAUC,<br>temporal<br>validation | Brier reduction,<br>development | Brier reduction,<br>temporal<br>validation |
| --- | --- | --- | --- | --- |
| **M1 vs M0 (+ body fat % and VAT)** |  |  |  |  |
| Logistic regression | 0.0209<br>(0.0024–0.0402) | 0.0209<br>(0.0032–0.0375) | 0.0081<br>(0.0006–0.0155) | 0.0069<br>(0.0004–0.0131) |
| Elastic-net | 0.0169<br>(–0.0019 to 0.0363) | 0.0129<br>(–0.0027 to 0.0275) | 0.0068<br>(0.0001–0.0135) | 0.0054<br>(0.0008–0.0099) |
| XGBoost | 0.0246<br>(0.0007–0.0488) | 0.0243<br>(0.0045–0.0433) | 0.0082<br>(–0.0007 to 0.0172) | 0.0106<br>(0.0020–0.0193) |
| **M2 vs M1 (+ ASMI)** |  |  |  |  |
| Logistic regression | –0.0030<br>(–0.0069 to 0.0011) | –0.0008<br>(–0.0036 to 0.0020) | –0.0018<br>(–0.0033 to –0.0003) | –0.0004<br>(–0.0013 to 0.0006) |
| Elastic-net | –0.0083<br>(–0.0187 to 0.0023) | –0.0017<br>(–0.0075 to 0.0039) | –0.0030<br>(–0.0072 to 0.0013) | –0.0003<br>(–0.0021 to 0.0015) |
| XGBoost | 0.0092<br>(–0.0044 to 0.0229) | –0.0112<br>(–0.0218 to –0.0007) | 0.0049<br>(–0.0002 to 0.0099) | –0.0033<br>(–0.0073 to 0.0009) |
| **M3A vs M2 (+ absolute grip strength;**<br>**primary)** |  |  |  |  |
| **Logistic regression** | **–0.0046**<br>(–0.0107 to 0.0013) | **0.0045**<br>(–0.0009 to 0.0098) | **–0.0013**<br>(–0.0035 to 0.0010) | **0.0018**<br>(–0.0002 to 0.0037) |
| Elastic-net | 0.0001<br>(–0.0057 to 0.0063) | –0.0004<br>(–0.0009 to 0.0001) | –0.0002<br>(–0.0021 to 0.0019) | –0.0001<br>(–0.0002 to –0.0001) |
| XGBoost | 0.0016<br>(–0.0065 to 0.0093) | 0.0012<br>(–0.0013 to 0.0037) | –0.0002<br>(–0.0031 to 0.0026) | 0.0002<br>(–0.0005 to 0.0010) |
| **M3B vs M2 (+ relative grip strength;**<br>**sensitivity)** |  |  |  |  |
| Logistic regression | –0.0022<br>(–0.0080 to 0.0030) | 0.0029<br>(–0.0014 to 0.0074) | –0.0007<br>(–0.0028 to 0.0014) | 0.0014<br>(–0.0003 to 0.0031) |
| Elastic-net | 0.0026<br>(–0.0017 to 0.0071) | 0.0018<br>(–0.0002 to 0.0040) | 0.0009<br>(–0.0005 to 0.0023) | 0.0008<br>(0.0002–0.0013) |
| XGBoost | –0.0052<br>(–0.0151 to 0.0038) | 0.0039<br>(–0.0036 to 0.0113) | –0.0022<br>(–0.0060 to 0.0014) | 0.0008<br>(–0.0018 to 0.0034) |

ΔAUC is the extended minus the reference model; Brier reduction is the reference minus the extended model, so positive values indicate improvement for both. Estimates are paired on the same participants, with 95% CIs from 2000 stratified paired bootstrap resamples (development n=508; temporal validation n=616). The prespecified primary comparison (M3A vs M2, logistic regression) is shown in bold. No adjustment was made for multiplicity. ASMI=appendicular skeletal muscle index. AUC=area under the receiver operating characteristic curve. VAT=visceral adipose tissue.

For the primary comparison (M3A vs M2 with logistic regression), ΔAUC was −0.0046 (−0.0107 to 0.0013) in development and 0.0045 (−0.0009 to 0.0098) in validation. In validation, the changes in average precision (0.0041 [−0.0024 to 0.0107]), Brier score (reduction 0.0018 [−0.0002 to 0.0037]) and log loss (reduction 0.0040 [−0.0004 to 0.0084]) were small and their intervals included zero. Across the 12 grip-strength comparisons (two definitions, three algorithms and two cohorts), ΔAUC ranged from −0.0052 to 0.0045 and every 95% CI included zero. Among grip-strength comparisons, the only intervals that excluded zero concerned very small changes in the Brier score (less than 0.001) and log loss (less than 0.002) with elastic-net in validation (Supplementary Table S3).

### 3.4. Calibration and clinical utility

In the development cohort, OOF predictions were well calibrated in the large (CITL −0.01 to 0.00 across the logistic ladder), with calibration slopes of 0.88–0.95. In validation, all logistic models slightly overestimated risk (CITL −0.15 to −0.12) and had slopes below 1 (0.80–0.84), consistent with the lower outcome prevalence. M3A and M2 had almost identical calibration (CITL −0.13 [−0.22 to −0.04] vs −0.14 [−0.24 to −0.05]; slope 0.81 [0.65–1.00] vs 0.80 [0.64–0.99]; Fig. 4A), and their decision curves were nearly superimposed (Fig. 4B). The incremental net benefit of M3A over M2 was positive at 32 of 46 thresholds, but its 95% CI excluded zero at only seven non-contiguous thresholds (0.13, 0.17, 0.18, 0.32, 0.38, 0.47 and 0.48; Fig. 4C). In development, it was positive at 17 thresholds; its CI never lay entirely above zero and lay entirely below zero at one threshold (0.43) (Supplementary Fig. S1).

**Fig. 4.**
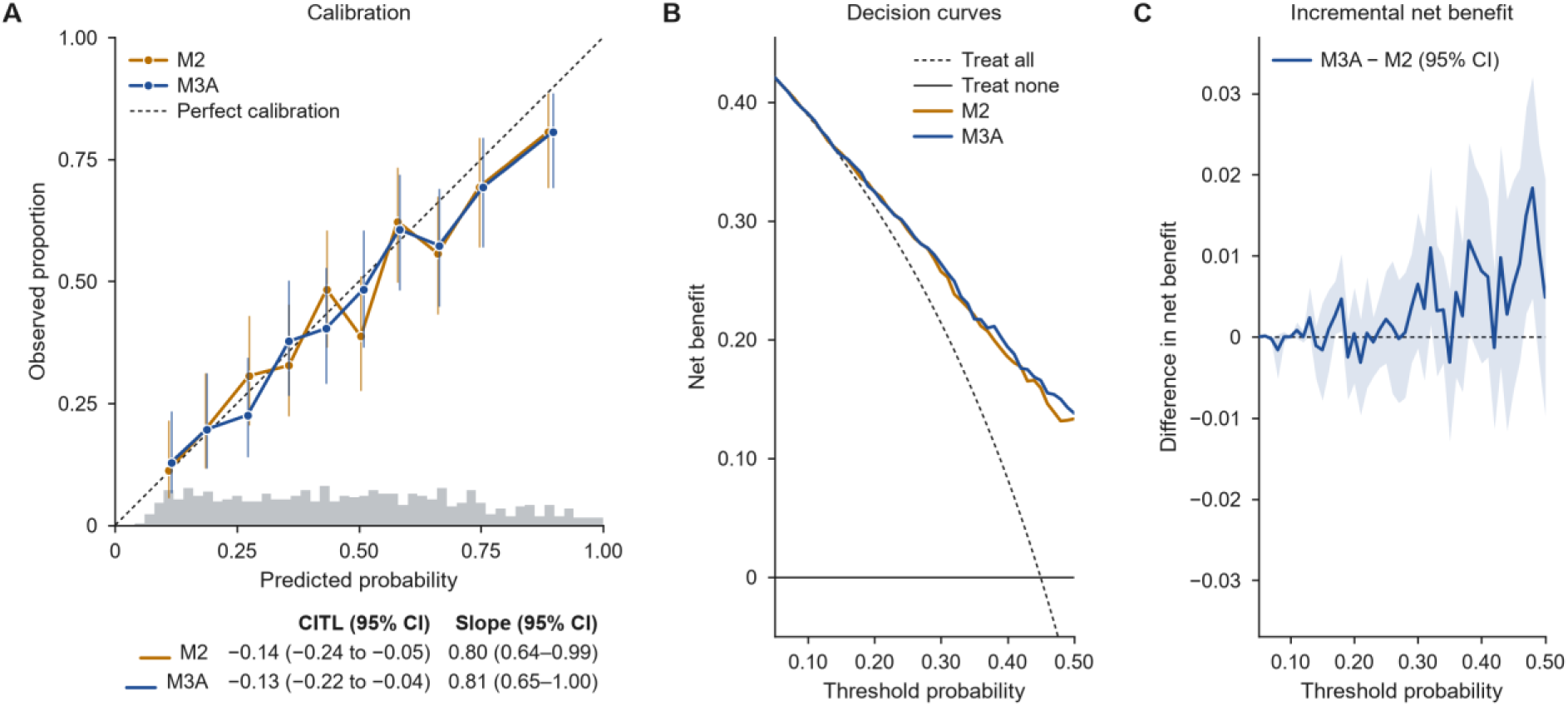
Calibration and decision curves for the primary comparison in temporal validation. Logistic regression M2 and M3A in the temporal validation cohort (n=616). (A) Observed proportions against mean predicted probabilities in ten equal-size groups, with Wilson 95% CIs; the dashed line shows perfect calibration, and the grey histogram shows the distribution of M3A predictions. CITL (intercept with the slope fixed at 1) and the calibration slope are shown with 2000-resample bootstrap 95% CIs. (B) Net benefit across the prespecified threshold range of 0.05–0.50; the dashed and solid black lines show the treat-all and treat-none strategies. (C) Difference in net benefit between M3A and M2, with the 95% CI band from the same paired bootstrap resamples. Predictions were not recalibrated. CITL=calibration-in-the-large.

### 3.5. Survey-weighted sensitivity analysis

With survey weights, AUCs were higher in development (for example, 0.803 [0.744–0.851] for M2) and similar in validation (0.757 [0.708–0.807] for M2). The adiposity increment was of similar size (weighted ΔAUC 0.0266 [−0.0043 to 0.0581] in development and 0.0275 [0.0063–0.0517] in validation), although the design-based interval included zero in development. The primary grip-strength increment remained close to zero (−0.0080 [−0.0206 to 0.0029] and 0.0007 [−0.0061 to 0.0080]), and in development adding grip strength slightly worsened the weighted Brier score (−0.0037 [−0.0073 to −0.0003]) (Supplementary Fig. S3; Supplementary Table S4).

### 3.6. Model explanations

In the development cohort, logistic regression M3A ranked absolute grip strength fourth of eight predictors by mean absolute cross-fitted contribution (0.192 log-odds; 95% CI 0.177–0.206), after VAT mass (0.622), body fat percentage (0.351) and ASMI (0.310) (Fig. 5B). Its standardised coefficient was negative in four of five outer folds (mean −0.226; range −0.354 to 0.011; Fig. 5A). XGBoost ranked grip strength last (mean absolute TreeSHAP value 0.0184; 95% CI 0.0172–0.0196), and its attributions were small and separated by sex (Fig. 5C and D). Relative grip strength ranked fourth with logistic regression and fifth with XGBoost. The models therefore used grip strength, but this did not translate into better discrimination, calibration or net benefit.

**Fig. 5.**
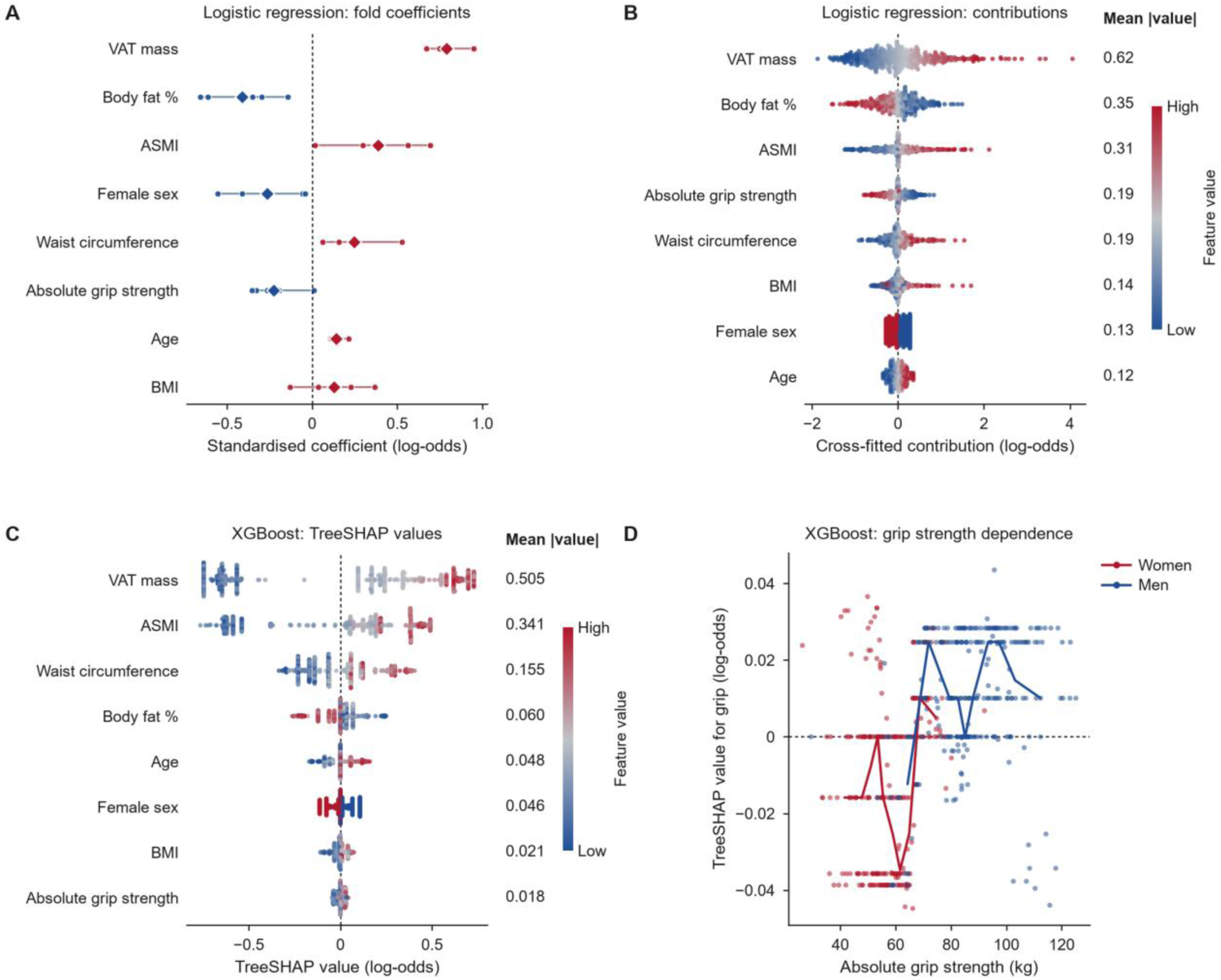
Cross-fitted explanations of model M3A in the development cohort. All values were obtained from outer-fold models that had not been trained on the participant concerned (n=508). (A) Standardised logistic regression coefficients in the five outer folds (dots) and their mean (diamonds); red indicates a positive and blue a negative mean coefficient; the coefficient for sex compares women with men. (B) Cross-fitted logistic regression contributions (log-odds) and (C) XGBoost TreeSHAP values, with features ordered by mean absolute value (right-hand column); colours show each feature’s value scaled between its 5th and 95th percentiles (women are coded as high). (D) TreeSHAP values for absolute grip strength by sex; lines join the medians of ten quantile bins in each sex. Attributions describe model behaviour and do not indicate incremental value or causal effects.

## 4. Discussion

In two consecutive NHANES cycles, with a prespecified feature ladder and frozen models, DXA-measured adiposity added a small but reproducible increment to anthropometry for concurrent cardiometabolic risk stratification in middle-aged adults, whereas neither muscle mass nor grip strength added further. For the primary comparison, the grip-strength increment was slightly negative in development and slightly positive in temporal validation, and both intervals included zero. The same pattern held for relative grip strength, for elastic-net and XGBoost, and in survey-weighted analyses; calibration was unchanged and the incremental net benefit was not consistently positive. These findings place an upper bound on the added value of grip strength in this setting: any improvement beyond detailed body composition is likely to be small, with an upper confidence limit for ΔAUC of 0.010 in temporal validation.

Grip strength is consistently associated with cardiovascular outcomes and mortality [6,7], and lower grip strength normalised to body mass has been associated with diabetes, dyslipidaemia and hypertension in NHANES 2011–2014, the same cycles analysed here [31]. Cross-sectional studies have linked low relative grip strength to the metabolic syndrome, although in one study the association was attenuated after adjustment for BMI [10,11]. Where incremental value has been tested, the gains have been modest: in UK Biobank, adding grip strength increased the C-index of established cardiovascular risk scores by 0.002–0.005 [8], and in older Finnish adults normalised grip strength improved the prediction of type 2 diabetes, mainly in women [9]. Conversely, in older adults in Hong Kong, grip strength and gait speed improved the prediction of adverse outcomes beyond a sarcopenia screening questionnaire, whereas DXA muscle mass added little further [32]; the value of each component therefore depends on what the reference model already contains. Our results are consistent with these reports and extend them in two ways. First, the reference model already contained DXA adiposity and appendicular muscle mass, which is the information that grip strength is often thought to approximate. Second, the comparison was prespecified, paired and repeated in a later survey cycle with frozen models. An association between grip strength and cardiometabolic status and a lack of incremental discrimination are therefore not contradictory [12].

The explanation analyses help to reconcile these observations. Logistic regression gave grip strength a mid-ranked attribution, XGBoost placed it last, and removing it barely changed performance in either case. Attribution methods describe how a fitted model distributes a prediction among correlated inputs [26]; they do not show that an input carries information that the others lack. Absolute grip strength is strongly structured by sex and body size, which the reference model already represented through sex, BMI, waist circumference and ASMI. Its conditional coefficient was mostly negative (greater strength, lower risk for a given body composition) but changed sign in one fold. Non-zero attribution without incremental value is what would be expected from a correlated but largely redundant predictor.

Participants with the outcome had higher absolute but lower relative grip strength, which reflects the larger proportion of men and the larger body size in this group. Normalising grip strength by body weight removes part of this size dependence but ties the measure to its denominator, which is itself associated with cardiometabolic risk [31]. Neither definition added information once BMI, VAT and ASMI were in the model, which suggests that the cardiometabolic information carried by grip strength overlaps substantially with body composition. This interpretation is consistent with the observation in older men that sarcopenic obesity did not confer greater metabolic risk than obesity alone [5].

The adiposity increment of about 0.02 in AUC was small, but it was reproduced in both cycles and was accompanied by an improvement in the Brier score. VAT is an established correlate of metabolic risk [2,3,33], and it received the largest attribution in the fitted models. In a companion analysis of the next two NHANES cycles (2015–2018; 562 development and 428 validation participants aged 40–59 years; manuscript in preparation), with the same outcome definition and the same baseline predictors, adding body fat percentage and VAT increased the temporal-validation AUC by 0.0569 (95% CI 0.0276–0.0882), and most of this increment was attributable to VAT. Because those cycles do not overlap with the present samples, the adiposity increment was observed in independent NHANES samples from 2011 to 2018. This supports DXA adiposity, rather than muscle function, as the more informative addition to anthropometry for this outcome. The companion results are reported separately and were not used in any part of the present analysis.

Where DXA is available, these results do not support adding grip strength solely to improve the statistical stratification of concurrent cardiometabolic risk. The question is different where DXA is not available: grip strength is cheap and portable and might partly substitute for body composition. That hypothesis requires a separately specified model without DXA predictors and cannot be inferred from the present full models. Muscle function may also matter for outcomes not studied here, such as incident disease, disability and mortality [6,7,34]; our findings concern concurrent phenotype stratification only.

The study has several strengths. Waist circumference was excluded from the outcome because it was also a predictor, predictors entered only through a whitelist, and all imputation, scaling, encoding and tuning took place within training folds. The analysis plan was frozen before training, the primary comparison was prespecified, and all comparisons were paired on the same participants. Temporal validation used frozen models, the access history of the validation data is fully disclosed, and model explanations were cross-fitted and kept separate from the evaluation of incremental value. A survey-weighted sensitivity analysis with design-based intervals was also performed.

The study also has limitations. First, the design was cross-sectional and cannot address causality or future events. Second, the samples were modest; the ΔAUC intervals were narrow in absolute terms (about ±0.005 for the primary comparison), but small increments cannot be excluded, and larger development samples would be needed for more complex models [35]. Third, the complete-case requirements for DXA and the fasting subsample excluded most age-eligible participants, which may have introduced selection bias; weighting addresses representativeness but not this restriction. Fourth, the age range was limited to 40–59 years by the DXA protocol, and performance was not examined separately by sex or by race and ethnicity, whose composition differed between the cohorts. Fifth, grip strength is only one aspect of muscle function; gait speed, chair stands and lower-limb strength were not examined. Sixth, the validation cohort was described before it was locked, read three times during scoring and opened again afterwards for descriptive purposes; no model or threshold was changed, but it is not an untouched test set, and external validation in other populations remains necessary [36]. Seventh, the bootstrap of fixed predictions does not capture model-fitting uncertainty, and the comparisons were not adjusted for multiplicity. Finally, the outcome is an operational definition that depends partly on medication use, and a standard five-component metabolic syndrome outcome was not analysed.

## 5. Conclusions

In middle-aged NHANES participants, DXA-derived adiposity improved concurrent cardiometabolic risk stratification beyond conventional anthropometry in both development and temporal validation, whereas grip strength, whether absolute or relative, did not add reproducible information beyond detailed body composition. Grip strength influenced individual predictions without improving discrimination, calibration or net benefit. Future work should evaluate grip strength in low-cost models without DXA and for prospective outcomes in independent cohorts.

## Data availability

NHANES public-use data are available from the National Center for Health Statistics (https://wwwn.cdc.gov/nchs/nhanes/). The analysis code and aggregate results are available from the corresponding author on reasonable request.

## Declaration of generative AI and AI-assisted technologies in the manuscript preparation process

During the preparation of this work the authors used Claude (Anthropic) and ChatGPT/Codex (OpenAI) to assist with analysis code, figure generation, drafting and translation. After using these tools, the authors reviewed and edited the content as needed and take full responsibility for the content of the published article.

## Acknowledgements

We thank the NHANES participants and the staff of the National Center for Health Statistics.

## Abbreviations

ASMI: appendicular skeletal muscle index
AUC: area under the receiver operating characteristic curve
BMI: body mass index
CI: confidence interval
CITL: calibration-in-the-large
DXA: dual-energy X-ray absorptiometry
HDL: high-density lipoprotein
NHANES: National Health and Nutrition Examination Survey
OOF: out-of-fold
PSU: primary sampling unit
SHAP: Shapley additive explanations
SMD: standardised mean difference
VAT: visceral adipose tissue

