## Supplementary Material for "Does handgrip strength improve cardiometabolic risk stratification beyond detailed body composition? NHANES 2011–2014 with temporal validation"

### Contents

Supplementary Methods S1. Variable definitions
Supplementary Methods S2. Access history of the temporal validation data
Supplementary Methods S3. Survey-weighted sensitivity analysis
Supplementary Table S1. Characteristics by outcome status within each cohort
Supplementary Table S2. Performance of all 15 models
Supplementary Table S3. Paired increments for all comparisons, algorithms and cohorts
Supplementary Table S4. Survey-weighted sensitivity analysis
Supplementary Table S5. Hyperparameters
Supplementary Table S6. TRIPOD+AI checklist
Supplementary Fig. S1. Calibration and decision curves in the development cohort
Supplementary Fig. S2. AUC of all 15 models
Supplementary Fig. S3. Survey-weighted change in AUC

### Supplementary Methods S1. Variable definitions

Suffixes _G and _H denote the 2011–2012 and 2013–2014 cycles. Values outside the plausibility ranges were set to missing. Only the predictors listed in Section 2.4 entered the models; all other variables were used for eligibility, outcome definition, description or weighting.

| Variable | NHANES file | NHANES variable(s) | Definition and processing |
|---|---|---|---|
| Age | DEMO_G / DEMO_H | RIDAGEYR | Age at screening, years; primary analysis restricted to 40–59 |
| Sex | DEMO_G / DEMO_H | RIAGENDR | Male or female |
| Race and ethnicity | DEMO_G / DEMO_H | RIDRETH3 | Six categories (descriptive only; not a predictor) |
| Education; income-to-poverty ratio | DEMO_G / DEMO_H | DMDEDUC2; INDFMPIR | Descriptive only |
| Pregnancy | DEMO; UCPREG | RIDEXPRG; URXPREG | Excluded if pregnant at examination or positive urine pregnancy test |
| Height, weight, BMI, waist circumference | BMX_G / BMX_H | BMXHT, BMXWT, BMXBMI, BMXWAIST | Plausibility ranges: height 100–250 cm, weight 20–350 kg, BMI 10–100 kg/m², waist 40–250 cm |
| Body fat percentage | DXX_G / DXX_H | DXDTOPF; DXAEXSTS | Whole-body fat %, valid scans only (DXAEXSTS=1); range 1–75% |
| Appendicular lean mass; ASMI | DXX_G / DXX_H | DXDLALE, DXDRALE, DXDLLLE, DXDRLLE | Sum of arm and leg lean mass excluding bone mineral content (kg); ASMI=ALM/height² (kg/m²) |
| VAT mass | DXXAG_G / DXXAG_H | DXXVFATM; DXXAGST | Visceral adipose tissue mass in the android region (g), valid only if DXXAGST=1; range 1–5000 g |
| Grip strength | MGX_G / MGX_H | MGDCGSZ; MGDEXSTS | Combined grip strength: sum of the largest reading from each hand (kg); complete test only; range 1–160 kg |
| Relative grip strength | MGX; BMX | MGDCGSZ/BMXWT | Combined grip strength divided by body weight (kg/kg) |
| Blood pressure | BPX_G / BPX_H | BPXSY1–4; BPXDI1–4 | Mean of readings 2–4 when available, otherwise mean of all readings |
| Antihypertensive medication | BPQ_G / BPQ_H | BPQ050A | Currently taking prescribed medicine for hypertension |
| Fasting glucose; fasting time | GLU_G / GLU_H | LBXGLU; PHAFSTHR | Plasma glucose (mg/dL); fasting duration 8–24 h required |
| Triglycerides; fasting weight | TRIGLY_G / TRIGLY_H | LBXTR; WTSAF2YR | Triglycerides (mg/dL); fasting-subsample 2-year weight |
| HDL cholesterol | HDL_G / HDL_H | LBDHDD | Direct HDL cholesterol (mg/dL) |
| Diabetes and treatment | DIQ_G / DIQ_H | DIQ010; DIQ050; DIQ070 | Doctor-diagnosed diabetes; insulin use; diabetic pill use |
| Survey design | DEMO_G / DEMO_H | SDMVSTRA; SDMVPSU | Masked variance strata and pseudo-PSUs (weighted sensitivity analysis only) |

### Supplementary Methods S2. Access history of the temporal validation data

Times are China Standard Time (UTC+8). All access events were logged; lock, authorisation and access records with SHA-256 hashes are archived with the analysis code.

| Date and time | Event | Use of validation data | Effect on models |
|---|---|---|---|
| 25 Aug 2026 | Data download, cleaning and cohort construction for both cycles | Outcome prevalence, predictor distributions, missingness and development–validation differences summarised for data-quality checks | None (no model fitted) |
| 26 Aug 2026 18:22 | Formal analysis plan frozen (SHA-256) | None | Outcome, predictors, ladder, algorithms, metrics fixed |
| 26 Aug 2026 18:45 | Validation files locked with file-system deny rules (57 | Access blocked | — |

| Date and time | Event | Use of validation data | Effect on models |
|---|---|---|---|
| | objects) | | |
| 26–27 Aug 2026 | Nested cross-validation, out-of-fold inference, explanations (development only) | None | — |
| 27 Aug 2026 00:27 | Final pipelines refitted on development data and frozen; validation plan frozen | None | Models and thresholds frozen |
| 27 Aug 2026 00:51–01:03 | One-time temporal validation authorised and run | Validation matrix read three times: two attempts stopped at calibration-diagnostic numerical checks before any output; third run completed and files relocked (01:02) | None (no refitting, tuning or recalibration) |
| 27 Aug 2026 01:07 | Final temporal-validation audit | None | — |
| 22 Sep 2026 00:28 | Authorised post-validation descriptive access (5 files; relocked after 3 s) | Descriptive characteristics, flow counts and survey design variables extracted; only aggregate results written | None (no predictions recomputed) |

### Supplementary Methods S3. Survey-weighted sensitivity analysis

The fasting-subsample 2-year examination weight (WTSAF2YR) was applied to the fixed predictions of the frozen models in each cohort. Weighted AUC was computed as the weighted Mann–Whitney statistic (ties counted as one half), and the weighted Brier score as the weighted mean squared error. Weighted calibration-in-the-large and slope were obtained from weighted logistic recalibration models.

Design-based 95% CIs were obtained with a Rao–Wu rescaling bootstrap: in each masked variance stratum with n pseudo-PSUs, n−1 PSUs were drawn with replacement and the weights of their members were multiplied by n/(n−1) times the number of times the PSU was drawn (2000 replicates, seed 20260922). The development cohort had 14 strata (3 with three PSUs) and the validation cohort 15 strata with two PSUs each. Paired increments used the same replicate weights for both models. As a check, the bootstrap standard error of the weighted outcome prevalence in the development cohort (0.036) agreed with Taylor linearisation (0.035); the corresponding values for mean BMI were 0.42 and 0.40.

The analysed samples are complete-case subsets of the fasting subsample, so weighted estimates correct for unequal selection probabilities but not for the complete-case restriction.

### Supplementary Table S1. Characteristics by outcome status within each cohort

| Characteristic | Development no high risk (n=259) | Development high risk (n=249) | SMD | Validation no high risk (n=339) | Validation high risk (n=277) | SMD |
|---|---|---|---|---|---|---|
| Age, years | 48.8 (5.7) | 49.9 (5.8) | 0.20 | 48.1 (5.7) | 50.0 (5.6) | 0.33 |
| Female sex | 155 (59.8) | 91 (36.5) | −0.48 | 186 (54.9) | 126 (45.5) | −0.19 |
| **Race and ethnicity** | | | | | | |
| Mexican American | 22 (8.5) | 30 (12.0) | 0.12 | 44 (13.0) | 42 (15.2) | 0.06 |
| Other Hispanic | 24 (9.3) | 25 (10.0) | 0.03 | 31 (9.1) | 32 (11.6) | 0.08 |
| Non-Hispanic White | 100 (38.6) | 97 (39.0) | 0.01 | 143 (42.2) | 117 (42.2) | 0.00 |
| Non-Hispanic Black | 70 (27.0) | 62 (24.9) | −0.05 | 56 (16.5) | 52 (18.8) | 0.06 |
| Non-Hispanic Asian | 35 (13.5) | 25 (10.0) | −0.11 | 53 (15.6) | 31 (11.2) | −0.13 |
| Other or multiracial | 8 (3.1) | 10 (4.0) | 0.05 | 12 (3.5) | 3 (1.1) | −0.16 |
| **Education** | | | | | | |
| Less than 9th grade | 14 (5.4) | 18 (7.2) | 0.08 | 18 (5.3) | 22 (7.9) | 0.11 |
| 9th–11th grade | 30 (11.6) | 34 (13.7) | 0.06 | 47 (13.9) | 40 (14.4) | 0.02 |
| High school or equivalent | 51 (19.7) | 54 (21.7) | 0.05 | 59 (17.4) | 61 (22.0) | 0.12 |
| Some college or associate degree | 71 (27.4) | 82 (32.9) | 0.12 | 98 (28.9) | 92 (33.2) | 0.09 |
| College graduate or above | 93 (35.9) | 61 (24.5) | −0.25 | 117 (34.5) | 62 (22.4) | −0.27 |
| Family income-to-poverty ratio | 2.87 (1.70) | 2.64 (1.68) | −0.14 | 2.94 (1.70) | 2.67 (1.67) | −0.16 |
| **Body size and composition** | | | | | | |
| BMI, kg/m² | 27.4 (5.7) | 31.7 (6.8) | 0.67 | 27.3 (5.7) | 31.5 (6.7) | 0.67 |
| Waist circumference, cm | 94.4 (13.3) | 106.6 (15.6) | 0.84 | 94.4 (13.3) | 105.0 (14.1) | 0.77 |
| Body fat, % | 34.1 (8.7) | 34.0 (8.1) | −0.02 | 33.4 (8.2) | 34.8 (8.0) | 0.18 |
| VAT mass, g | 480 (237) | 716 (275) | 0.92 | 492 (246) | 708 (278) | 0.82 |
| ASMI, kg/m² | 7.38 (1.46) | 8.58 (1.52) | 0.80 | 7.41 (1.45) | 8.33 (1.66) | 0.59 |
| **Muscle function** | | | | | | |
| Combined grip strength, kg | 69.6 (19.7) | 77.3 (19.2) | 0.39 | 72.6 (19.3) | 74.2 (21.5) | 0.08 |
| Grip strength/body weight, kg/kg | 0.93 (0.25) | 0.87 (0.23) | −0.23 | 0.97 (0.24) | 0.86 (0.25) | −0.43 |
| **Outcome components** | | | | | | |
| Elevated blood pressure | 58 (22.4) | 182 (73.1) | 1.18 | 60 (17.7) | 193 (69.7) | 1.23 |
| Elevated fasting glucose | 62 (23.9) | 213 (85.5) | 1.58 | 76 (22.4) | 234 (84.5) | 1.59 |
| Elevated triglycerides | 25 (9.7) | 134 (53.8) | 1.08 | 19 (5.6) | 139 (50.2) | 1.15 |
| Low HDL cholesterol | 15 (5.8) | 120 (48.2) | 1.09 | 30 (8.8) | 137 (49.5) | 1.00 |

| Characteristic | Development no high risk (n=259) | Development high risk (n=249) | SMD | Validation no high risk (n=339) | Validation high risk (n=277) | SMD |
|---|---|---|---|---|---|---|
| **Clinical measurements** | | | | | | |
| Systolic blood pressure, mmHg | 118.1 (13.3) | 127.5 (16.3) | 0.63 | 115.8 (13.2) | 127.6 (17.6) | 0.75 |
| Diastolic blood pressure, mmHg | 72.0 (9.2) | 76.6 (11.2) | 0.44 | 69.5 (9.2) | 73.7 (11.6) | 0.40 |
| Fasting glucose, mg/dL, median (IQR) | 95 (90–99) | 109 (102–126) | 0.88 | 96 (91–99) | 108 (101–122) | 0.80 |
| Triglycerides, mg/dL, median (IQR) | 89 (67–117) | 158 (101–208) | 0.77 | 83 (60–108) | 150 (99–213) | 0.96 |
| HDL cholesterol, mg/dL | 60.5 (15.8) | 45.9 (12.9) | −1.01 | 59.3 (14.7) | 47.0 (14.7) | −0.84 |
| Antihypertensive medication | 31 (12.0) | 103 (41.4) | 0.70 | 24 (7.1) | 117 (42.2) | 0.89 |
| Insulin or glucose-lowering pills | 5 (1.9) | 57 (22.9) | 0.67 | 6 (1.8) | 51 (18.4) | 0.57 |

Values are unweighted mean (SD), median (IQR) or n (%). SMD compares participants with and without high cardiometabolic risk within each cohort (high risk minus no high risk). Abbreviations as in Table 1.

### Supplementary Table S2. Performance of all 15 models in the development and temporal validation cohorts

| Algorithm and model | Cohort | AUC (95% CI) | Average precision (95% CI) | Brier score (95% CI) | Log loss | CITL (95% CI) | Slope (95% CI) |
|---|---|---|---|---|---|---|---|
| **Logistic regression** | | | | | | | |
| M0 | Development | 0.748 (0.704–0.789) | 0.735 (0.687–0.784) | 0.204 (0.188–0.220) | 0.593 | 0.00 (−0.09 to 0.09) | 0.95 (0.74–1.18) |
| | Validation | 0.722 (0.681–0.762) | 0.663 (0.616–0.716) | 0.213 (0.198–0.228) | 0.615 | −0.12 (−0.20 to −0.04) | 0.84 (0.65–1.04) |
| M1 | Development | 0.768 (0.727–0.809) | 0.752 (0.702–0.804) | 0.196 (0.179–0.212) | 0.575 | 0.00 (−0.10 to 0.10) | 0.93 (0.74–1.16) |
| | Validation | 0.743 (0.703–0.781) | 0.684 (0.634–0.736) | 0.206 (0.190–0.222) | 0.602 | −0.15 (−0.24 to −0.06) | 0.81 (0.64–1.00) |
| M2 | Development | 0.766 (0.725–0.807) | 0.749 (0.700–0.800) | 0.197 (0.180–0.214) | 0.579 | 0.00 (−0.10 to 0.10) | 0.90 (0.72–1.13) |
| | Validation | 0.742 (0.702–0.780) | 0.686 (0.636–0.737) | 0.206 (0.190–0.222) | 0.602 | −0.14 (−0.24 to −0.05) | 0.80 (0.64–0.99) |
| M3A | Development | 0.761 (0.719–0.802) | 0.744 (0.694–0.795) | 0.199 (0.181–0.216) | 0.582 | −0.01 (−0.11 to 0.09) | 0.88 (0.70–1.11) |
| | Validation | 0.746 (0.707–0.785) | 0.690 (0.640–0.741) | 0.204 (0.189–0.220) | 0.598 | −0.13 (−0.22 to −0.04) | 0.81 (0.65–1.00) |
| M3B | Development | 0.763 (0.722–0.805) | 0.746 (0.696–0.798) | 0.198 (0.180–0.215) | 0.581 | 0.00 (−0.11 to 0.09) | 0.89 (0.71–1.12) |
| | Validation | 0.745 (0.705–0.783) | 0.689 (0.639–0.740) | 0.205 (0.189–0.221) | 0.599 | −0.13 (−0.23 to −0.04) | 0.80 (0.64–1.00) |
| **Elastic-net** | | | | | | | |
| M0 | Development | 0.749 (0.705–0.789) | 0.732 (0.683–0.780) | 0.203 (0.189–0.219) | 0.593 | −0.01 (−0.09 to 0.07) | 1.05 (0.82–1.33) |
| | Validation | 0.729 (0.689–0.768) | 0.668 (0.620–0.720) | 0.211 (0.199–0.223) | 0.611 | −0.12 (−0.18 to −0.07) | 1.17 (0.91–1.46) |
| M1 | Development | 0.766 (0.726–0.807) | 0.750 (0.700–0.802) | 0.197 (0.181–0.212) | 0.578 | 0.00 (−0.09 to 0.08) | 1.03 (0.82–1.29) |
| | Validation | 0.742 (0.702–0.779) | 0.680 (0.630–0.733) | 0.206 (0.193–0.219) | 0.599 | −0.14 (−0.21 to −0.07) | 1.04 (0.82–1.29) |
| M2 | Development | 0.758 (0.716–0.798) | 0.747 (0.701–0.796) | 0.200 (0.186–0.214) | 0.581 | −0.01 (−0.08 to 0.07) | 1.20 (0.96–1.49) |
| | Validation | 0.740 (0.699–0.778) | 0.685 (0.636–0.736) | 0.206 (0.192–0.220) | 0.599 | −0.13 (−0.21 to −0.06) | 0.99 (0.78–1.22) |
| M3A | Development | 0.758 (0.715–0.799) | 0.746 (0.700–0.795) | 0.200 (0.186–0.214) | 0.582 | 0.00 (−0.08 to 0.07) | 1.17 (0.94–1.44) |
| | Validation | 0.740 (0.699–0.778) | 0.685 (0.638–0.736) | 0.206 (0.192–0.220) | 0.600 | −0.13 (−0.21 to −0.06) | 0.98 (0.78–1.22) |
| M3B | Development | 0.760 (0.718–0.802) | 0.751 (0.706–0.799) | 0.199 (0.185–0.212) | 0.579 | 0.00 (−0.08 to 0.07) | 1.26 (1.02–1.56) |
| | Validation | 0.742 (0.701–0.780) | 0.687 (0.638–0.738) | 0.205 (0.191–0.219) | 0.598 | −0.13 (−0.20 to −0.06) | 0.99 (0.79–1.23) |
| **XGBoost** | | | | | | | |
| M0 | Development | 0.737 (0.693–0.778) | 0.735 (0.688–0.781) | 0.208 (0.194–0.222) | 0.602 | 0.01 (−0.07 to 0.08) | 1.04 (0.82–1.27) |
| | Validation | 0.724 (0.683–0.764) | 0.667 (0.619–0.719) | 0.215 (0.206–0.224) | 0.620 | −0.12 (−0.16 to −0.08) | 1.51 (1.20–1.87) |
| M1 | Development | 0.761 (0.721–0.802) | 0.736 (0.685–0.789) | 0.199 (0.184–0.215) | 0.585 | 0.02 (−0.06 to 0.10) | 1.06 (0.85–1.29) |
| | Validation | 0.748 (0.710–0.785) | 0.689 (0.642–0.741) | 0.204 (0.189–0.220) | 0.595 | −0.12 (−0.21 to −0.04) | 0.91 (0.74–1.09) |
| M2 | Development | 0.771 (0.731–0.811) | 0.739 (0.689–0.793) | 0.195 (0.179–0.210) | 0.574 | 0.01 (−0.08 to 0.10) | 1.05 (0.85–1.28) |
| | Validation | 0.737 (0.698–0.775) | 0.685 (0.639–0.734) | 0.208 (0.192–0.223) | 0.603 | −0.12 (−0.21 to −0.03) | 0.82 (0.67–1.00) |

| Algorithm and model | Cohort | AUC (95% CI) | Average precision (95% CI) | Brier score (95% CI) | Log loss | CITL (95% CI) | Slope (95% CI) |
|---|---|---|---|---|---|---|---|
| M3A | Development | 0.772 (0.732–0.811) | 0.744 (0.695–0.797) | 0.195 (0.180–0.209) | 0.573 | 0.02 (−0.06 to 0.09) | 1.18 (0.97–1.42) |
| | Validation | 0.738 (0.699–0.776) | 0.688 (0.642–0.736) | 0.207 (0.192–0.223) | 0.602 | −0.12 (−0.20 to −0.03) | 0.82 (0.67–1.00) |
| M3B | Development | 0.765 (0.724–0.807) | 0.731 (0.681–0.787) | 0.197 (0.180–0.213) | 0.579 | 0.02 (−0.07 to 0.11) | 0.98 (0.79–1.21) |
| | Validation | 0.741 (0.702–0.779) | 0.685 (0.638–0.737) | 0.207 (0.190–0.223) | 0.602 | −0.09 (−0.19 to 0.00) | 0.79 (0.64–0.97) |

Development values are from out-of-fold predictions (n=508); validation values from frozen models scored once (n=616). 95% CIs from 2000 stratified bootstrap resamples. M0: age, sex, BMI, waist circumference; M1: M0 + body fat % and VAT; M2: M1 + ASMI; M3A: M2 + absolute grip strength; M3B: M2 + relative grip strength. CITL=calibration-in-the-large.

### Supplementary Table S3. Paired increments for all comparisons, algorithms and cohorts

| Comparison and algorithm | Cohort | ΔAUC (95% CI) | ΔAverage precision (95% CI) | Brier reduction (95% CI) | Log-loss reduction (95% CI) |
|---|---|---|---|---|---|
| **M1 vs M0 (+ body fat % and VAT)** | | | | | |
| Logistic regression | Development | 0.0209 (0.0024–0.0402) | 0.0162 (−0.0127 to 0.0447) | 0.0081 (0.0006–0.0155) | 0.0173 (−0.0037 to 0.0364) |
| | Validation | 0.0209 (0.0032–0.0375) | 0.0212 (−0.0032 to 0.0466) | 0.0069 (0.0004–0.0131) | 0.0129 (−0.0036 to 0.0287) |
| Elastic-net | Development | 0.0169 (−0.0019 to 0.0363) | 0.0174 (−0.0089 to 0.0450) | 0.0068 (0.0001–0.0135) | 0.0152 (−0.0023 to 0.0324) |
| | Validation | 0.0129 (−0.0027 to 0.0275) | 0.0120 (−0.0098 to 0.0353) | 0.0054 (0.0008–0.0099) | 0.0115 (0.0005–0.0223) |
| XGBoost | Development | 0.0246 (0.0007–0.0488) | 0.0013 (−0.0371 to 0.0395) | 0.0082 (−0.0007 to 0.0172) | 0.0177 (−0.0018 to 0.0381) |
| | Validation | 0.0243 (0.0045–0.0433) | 0.0220 (−0.0053 to 0.0498) | 0.0106 (0.0020–0.0193) | 0.0254 (0.0052–0.0454) |
| **M2 vs M1 (+ ASMI)** | | | | | |
| Logistic regression | Development | −0.0030 (−0.0069 to 0.0011) | −0.0029 (−0.0078 to 0.0020) | −0.0018 (−0.0033 to −0.0003) | −0.0040 (−0.0075 to −0.0003) |
| | Validation | −0.0008 (−0.0036 to 0.0020) | 0.0016 (−0.0038 to 0.0064) | −0.0004 (−0.0013 to 0.0006) | −0.0006 (−0.0029 to 0.0019) |
| Elastic-net | Development | −0.0083 (−0.0187 to 0.0023) | −0.0026 (−0.0191 to 0.0139) | −0.0030 (−0.0072 to 0.0013) | −0.0038 (−0.0163 to 0.0096) |
| | Validation | −0.0017 (−0.0075 to 0.0039) | 0.0049 (−0.0036 to 0.0120) | −0.0003 (−0.0021 to 0.0015) | −0.0002 (−0.0042 to 0.0039) |
| XGBoost | Development | 0.0092 (−0.0044 to 0.0229) | 0.0033 (−0.0160 to 0.0221) | 0.0049 (−0.0002 to 0.0099) | 0.0106 (−0.0013 to 0.0224) |
| | Validation | −0.0112 (−0.0218 to −0.0007) | −0.0040 (−0.0242 to 0.0142) | −0.0033 (−0.0073 to 0.0009) | −0.0082 (−0.0176 to 0.0018) |
| **M3A vs M2 (+ absolute grip strength; primary)** | | | | | |
| Logistic regression | Development | −0.0046 (−0.0107 to 0.0013) | −0.0048 (−0.0118 to 0.0024) | −0.0013 (−0.0035 to 0.0010) | −0.0029 (−0.0079 to 0.0023) |
| | Validation | 0.0045 (−0.0009 to 0.0098) | 0.0041 (−0.0024 to 0.0107) | 0.0018 (−0.0002 to 0.0037) | 0.0040 (−0.0004 to 0.0084) |
| Elastic-net | Development | 0.0001 (−0.0057 to 0.0063) | −0.0007 (−0.0097 to 0.0072) | −0.0002 (−0.0021 to 0.0019) | −0.0003 (−0.0050 to 0.0046) |
| | Validation | −0.0004 (−0.0009 to 0.0001) | 0.0002 (−0.0007 to 0.0019) | −0.0001 (−0.0002 to −0.0001) | −0.0003 (−0.0004 to −0.0001) |
| XGBoost | Development | 0.0016 (−0.0065 to 0.0093) | 0.0044 (−0.0113 to 0.0203) | −0.0002 (−0.0031 to 0.0026) | 0.0006 (−0.0070 to 0.0079) |
| | Validation | 0.0012 (−0.0013 to 0.0037) | 0.0027 (−0.0023 to 0.0080) | 0.0002 (−0.0005 to 0.0010) | 0.0005 (−0.0013 to 0.0023) |
| **M3B vs M2 (+ relative grip strength; sensitivity)** | | | | | |
| Logistic regression | Development | −0.0022 (−0.0080 to 0.0030) | −0.0028 (−0.0087 to 0.0030) | −0.0007 (−0.0028 to 0.0014) | −0.0017 (−0.0066 to 0.0031) |
| | Validation | 0.0029 (−0.0014 to 0.0074) | 0.0027 (−0.0025 to 0.0081) | 0.0014 (−0.0003 to 0.0031) | 0.0033 (−0.0005 to 0.0072) |
| Elastic-net | Development | 0.0026 (−0.0017 to 0.0071) | 0.0035 (−0.0026 to 0.0099) | 0.0009 (−0.0005 to 0.0023) | 0.0019 (−0.0018 to 0.0062) |
| | Validation | 0.0018 (−0.0002 to 0.0040) | 0.0014 (−0.0013 to 0.0042) | 0.0008 (0.0002–0.0013) | 0.0017 (0.0004–0.0029) |
| XGBoost | Development | −0.0052 (−0.0151 to 0.0038) | −0.0083 (−0.0220 to 0.0057) | −0.0022 (−0.0060 to 0.0014) | −0.0054 (−0.0148 to 0.0036) |
| | Validation | 0.0039 (−0.0036 to 0.0113) | −0.0005 (−0.0158 to 0.0155) | 0.0008 (−0.0018 to 0.0034) | 0.0008 (−0.0058 to 0.0079) |

Increments are extended minus reference for AUC and average precision, and reference minus extended for Brier score and log loss (positive values indicate improvement). 95% CIs from 2000 stratified paired bootstrap resamples. No adjustment for multiplicity.

### Supplementary Table S4. Survey-weighted sensitivity analysis

(a) Weighted discrimination of all models

| Algorithm and model | Development unweighted AUC | Development weighted AUC (95% CI) | Validation unweighted AUC | Validation weighted AUC (95% CI) |
|---|---|---|---|---|
| **Logistic regression** | | | | |
| M0 | 0.748 | 0.777 (0.721–0.827) | 0.722 | 0.731 (0.670–0.789) |
| M1 | 0.768 | 0.804 (0.746–0.850) | 0.743 | 0.759 (0.709–0.809) |
| M2 | 0.766 | 0.803 (0.744–0.851) | 0.742 | 0.757 (0.708–0.807) |
| M3A | 0.761 | 0.795 (0.730–0.845) | 0.746 | 0.758 (0.710–0.805) |
| M3B | 0.763 | 0.797 (0.734–0.846) | 0.745 | 0.757 (0.710–0.805) |
| **Elastic-net** | | | | |
| M0 | 0.749 | 0.780 (0.727–0.826) | 0.729 | 0.737 (0.678–0.793) |
| M1 | 0.766 | 0.800 (0.745–0.846) | 0.742 | 0.756 (0.705–0.809) |
| M2 | 0.758 | 0.796 (0.732–0.845) | 0.740 | 0.752 (0.701–0.804) |
| M3A | 0.758 | 0.791 (0.723–0.843) | 0.740 | 0.752 (0.701–0.803) |
| M3B | 0.760 | 0.795 (0.728–0.846) | 0.742 | 0.754 (0.703–0.804) |
| **XGBoost** | | | | |
| M0 | 0.737 | 0.763 (0.713–0.806) | 0.724 | 0.735 (0.679–0.789) |
| M1 | 0.761 | 0.791 (0.733–0.836) | 0.748 | 0.761 (0.714–0.806) |
| M2 | 0.771 | 0.801 (0.733–0.855) | 0.737 | 0.748 (0.697–0.797) |
| M3A | 0.772 | 0.804 (0.734–0.857) | 0.738 | 0.750 (0.700–0.799) |
| M3B | 0.765 | 0.788 (0.719–0.845) | 0.741 | 0.754 (0.703–0.804) |

(b) Weighted paired increments

| Comparison and algorithm | Weighted ΔAUC, development | Weighted ΔAUC, validation | Weighted Brier reduction, development | Weighted Brier reduction, validation |
|---|---|---|---|---|
| **M1 vs M0 (+ body fat % and VAT)** | | | | |
| Logistic regression | 0.0266 (−0.0043 to 0.0581) | 0.0275 (0.0063–0.0517) | 0.0101 (−0.0015 to 0.0223) | 0.0095 (0.0015–0.0178) |
| Elastic-net | 0.0207 (−0.0123 to 0.0555) | 0.0190 (0.0042–0.0339) | 0.0091 (−0.0020 to 0.0209) | 0.0072 (0.0027–0.0119) |
| XGBoost | 0.0275 (−0.0141 to 0.0670) | 0.0265 (0.0077–0.0452) | 0.0087 (−0.0050 to 0.0218) | 0.0130 (0.0051–0.0215) |
| **M2 vs M1 (+ ASMI)** | | | | |
| Logistic regression | −0.0010 (−0.0053 to 0.0039) | −0.0016 (−0.0049 to 0.0012) | −0.0007 (−0.0023 to 0.0007) | −0.0004 (−0.0014 to 0.0005) |
| Elastic-net | −0.0046 (−0.0214 to 0.0083) | −0.0038 (−0.0088 to 0.0012) | −0.0032 (−0.0079 to 0.0003) | −0.0003 (−0.0018 to 0.0010) |
| XGBoost | 0.0103 (−0.0104 to 0.0284) | −0.0133 (−0.0258 to −0.0006) | 0.0070 (−0.0017 to 0.0143) | −0.0048 (−0.0093 to 0.0000) |
| **M3A vs M2 (+ absolute grip strength; primary)** | | | | |
| Logistic regression | −0.0080 (−0.0206 to 0.0029) | 0.0007 (−0.0061 to 0.0080) | −0.0037 (−0.0073 to −0.0003) | 0.0011 (−0.0014 to 0.0035) |
| Elastic-net | −0.0049 (−0.0141 to 0.0037) | −0.0002 (−0.0008 to 0.0004) | −0.0023 (−0.0054 to 0.0005) | −0.0001 (−0.0002 to 0.0000) |

| Comparison and algorithm | Weighted ΔAUC, development | Weighted ΔAUC, validation | Weighted Brier reduction, development | Weighted Brier reduction, validation |
|---|---|---|---|---|
| XGBoost | 0.0030 (−0.0114 to 0.0126) | 0.0022 (−0.0010 to 0.0050) | −0.0010 (−0.0046 to 0.0022) | 0.0004 (−0.0005 to 0.0014) |
| **M3B vs M2 (+ relative grip strength; sensitivity)** | | | | |
| Logistic regression | −0.0059 (−0.0152 to 0.0026) | 0.0001 (−0.0049 to 0.0052) | −0.0032 (−0.0061 to −0.0006) | 0.0009 (−0.0013 to 0.0031) |
| Elastic-net | −0.0011 (−0.0075 to 0.0060) | 0.0017 (−0.0014 to 0.0046) | −0.0010 (−0.0021 to 0.0003) | 0.0006 (−0.0002 to 0.0013) |
| XGBoost | −0.0130 (−0.0250 to −0.0020) | 0.0064 (−0.0017 to 0.0143) | −0.0044 (−0.0097 to 0.0002) | 0.0018 (−0.0011 to 0.0046) |

Weights: fasting-subsample 2-year examination weights. 95% CIs from a Rao–Wu rescaling bootstrap (2000 replicates) resampling pseudo-PSUs within masked variance strata (Supplementary Methods S3). Survey-weighted outcome prevalence: 45.9% (95% CI 40.0–53.3) in development and 43.9% (37.6–51.1) in validation.

### Supplementary Table S5. Hyperparameter search spaces and hyperparameters of the frozen pipelines

Search spaces (random search inside training folds; 30 candidates for elastic-net and 40 for XGBoost; refit criterion AUC):

Elastic-net: C: 0.001, 0.003, 0.01, 0.03, 0.1, 0.3, 1.0, 3.0, 10.0, 30.0, 100.0; l1_ratio: 0.0, 0.1, 0.25, 0.5, 0.75, 0.9, 1.0

XGBoost: n_estimators: 100, 200, 300, 500; learning_rate: 0.01, 0.03, 0.05, 0.1; max_depth: 1, 2, 3; min_child_weight: 3, 5, 10, 20; subsample: 0.6, 0.75, 0.9, 1.0; colsample_bytree: 0.6, 0.75, 0.9, 1.0; gamma: 0.0, 0.1, 0.5, 1.0; reg_alpha: 0.0, 0.01, 0.1, 1.0; reg_lambda: 1.0, 5.0, 10.0, 20.0

| Feature set | Algorithm | Inner-CV AUC | Hyperparameters of the frozen pipeline |
|---|---|---|---|
| M0 | Logistic regression | — | Fixed: L2 penalty, C=1, lbfgs solver (no tuning) |
| M0 | Elastic-net | 0.747 | C=0.03; l1_ratio=0.0 |
| M0 | XGBoost | 0.746 | colsample_bytree=0.6; gamma=0.5; learning_rate=0.01; max_depth=2; min_child_weight=5; n_estimators=200; reg_alpha=1.0; reg_lambda=10.0; subsample=0.6 |
| M1 | Logistic regression | — | Fixed: L2 penalty, C=1, lbfgs solver (no tuning) |
| M1 | Elastic-net | 0.772 | C=0.03; l1_ratio=0.0 |
| M1 | XGBoost | 0.771 | colsample_bytree=0.75; gamma=0.1; learning_rate=0.05; max_depth=2; min_child_weight=20; n_estimators=100; reg_alpha=0.01; reg_lambda=1.0; subsample=0.75 |
| M2 | Logistic regression | — | Fixed: L2 penalty, C=1, lbfgs solver (no tuning) |
| M2 | Elastic-net | 0.765 | C=0.03; l1_ratio=0.0 |
| M2 | XGBoost | 0.774 | colsample_bytree=0.6; gamma=1.0; learning_rate=0.03; max_depth=1; min_child_weight=10; n_estimators=300; reg_alpha=0.0; reg_lambda=10.0; subsample=0.6 |
| M3A | Logistic regression | — | Fixed: L2 penalty, C=1, lbfgs solver (no tuning) |
| M3A | Elastic-net | 0.764 | C=0.03; l1_ratio=0.0 |
| M3A | XGBoost | 0.773 | colsample_bytree=0.6; gamma=1.0; learning_rate=0.03; max_depth=1; min_child_weight=10; n_estimators=300; reg_alpha=0.0; reg_lambda=10.0; subsample=0.6 |
| M3B | Logistic regression | — | Fixed: L2 penalty, C=1, lbfgs solver (no tuning) |
| M3B | Elastic-net | 0.766 | C=0.03; l1_ratio=0.0 |
| M3B | XGBoost | 0.775 | colsample_bytree=0.75; gamma=1.0; learning_rate=0.1; max_depth=1; min_child_weight=3; n_estimators=100; reg_alpha=1.0; reg_lambda=1.0; subsample=0.6 |

Inner-CV AUC is the mean AUC of the selected candidate across four inner folds of the final fit on all development participants. Logistic regression was not tuned. All pipelines include training-fold median imputation and standardisation of continuous predictors and mode imputation with one-hot encoding of sex.

### Supplementary Table S6. TRIPOD+AI checklist

| Item | Topic | Where reported |
|---|---|---|
| 1 | Title | Title page |
| 2 | Abstract | Structured abstract |
| 3a | Background: healthcare context and rationale | Introduction, paragraphs 1–3 |
| 3b | Target population and intended purpose | Introduction, paragraph 4; Section 2.1 (concurrent risk stratification in adults aged 40–59 years) |
| 3c | Known health inequalities | Table 1 (race and ethnicity); Discussion, limitations (performance not examined by sex or race and ethnicity) |
| 4 | Objectives | Introduction, paragraph 4 |
| 5a | Sources of data | Section 2.1 |
| 5b | Dates of data collection | Section 2.1 (NHANES 2011–2012 and 2013–2014 survey cycles) |
| 6a | Study setting | Section 2.1 (US general population survey) |
| 6b | Eligibility criteria | Section 2.2; Fig. 1A |
| 6c | Treatments | Section 2.3 (medication use contributes to outcome components and is excluded from predictors) |
| 7 | Data preparation | Sections 2.4–2.5; Supplementary Methods S1 |
| 8a | Outcome definition | Section 2.3 |
| 8b | Subjective outcome assessment | Not applicable (examination and laboratory measurements) |
| 8c | Blinding of outcome assessment | Outcome derived from survey measurements recorded independently of the models |
| 9a | Choice of initial predictors | Section 2.4 (prespecified feature ladder) |
| 9b | Predictor definitions | Section 2.4; Supplementary Methods S1 |
| 9c | Subjective predictor assessment | Not applicable |
| 10 | Sample size | Section 2.2 (all eligible participants); Discussion, limitations |
| 11 | Missing data | Sections 2.2 and 2.5 (training-fold median imputation); Section 3.1 |
| 12a | Data use | Sections 2.5–2.6 |
| 12b | Predictor handling | Section 2.5 |
| 12c | Model type and building steps | Section 2.5; Supplementary Table S5 |
| 12d | Heterogeneity | Not assessed across clusters; fold-to-fold coefficient variation in Section 3.6 |
| 12e | Performance measures | Section 2.7 |
| 12f | Model updating | None; predictions were not recalibrated (Section 2.6) |
| 12g | Calculation of predictions in evaluation | Section 2.6 (frozen pipelines applied once) |
| 13 | Class imbalance | Section 2.5 (no class-imbalance methods) |
| 14 | Fairness | Not assessed; Discussion, limitations |
| 15 | Model output | Section 2.5 (predicted probabilities) |
| 16 | Training versus evaluation data | Section 3.1; Table 1 (standardised mean differences) |
| 17 | Ethical approval | Section 2.10 |
| 18a | Funding | Declarations |
| 18b | Conflicts of interest | Declarations |
| 18c | Protocol | Section 2.1 (frozen analysis plan; shared with the code) |
| 18d | Registration | Section 2.1 (not registered) |
| 18e | Data sharing | Data availability statement |
| 18f | Code sharing | Data availability statement |
| 19 | Patient and public involvement | Section 2.10 (none) |
| 20a | Participant flow | Section 3.1; Fig. 1A |
| 20b | Participant characteristics | Table 1; Supplementary Table S1 |
| 20c | Comparison of development and evaluation data | Table 1 (standardised mean differences) |
| 21 | Participants and events in each analysis | Fig. 1A; Table 2 |
| 22 | Full model specification | Supplementary Table S5; frozen pipelines available (Data availability) |
| 23a | Performance with confidence intervals | Tables 2–3; Supplementary Tables S2–S4 |
| 23b | Heterogeneity in performance | Not examined |

| Item | Topic | Where reported |
|---|---|---|
| 24 | Model updating results | Not applicable (no updating) |
| 25 | Interpretation | Discussion |
| 26 | Limitations | Discussion, limitations |
| 27a | Handling of poor-quality or unavailable input data | Not applicable (research models, not intended for clinical deployment) |
| 27b | User interaction | Not applicable |
| 27c | Next steps for research | Discussion; Conclusions |

Checklist items from Collins et al., BMJ 2024;385:e078378 (Table 2). Section numbers refer to the main manuscript.

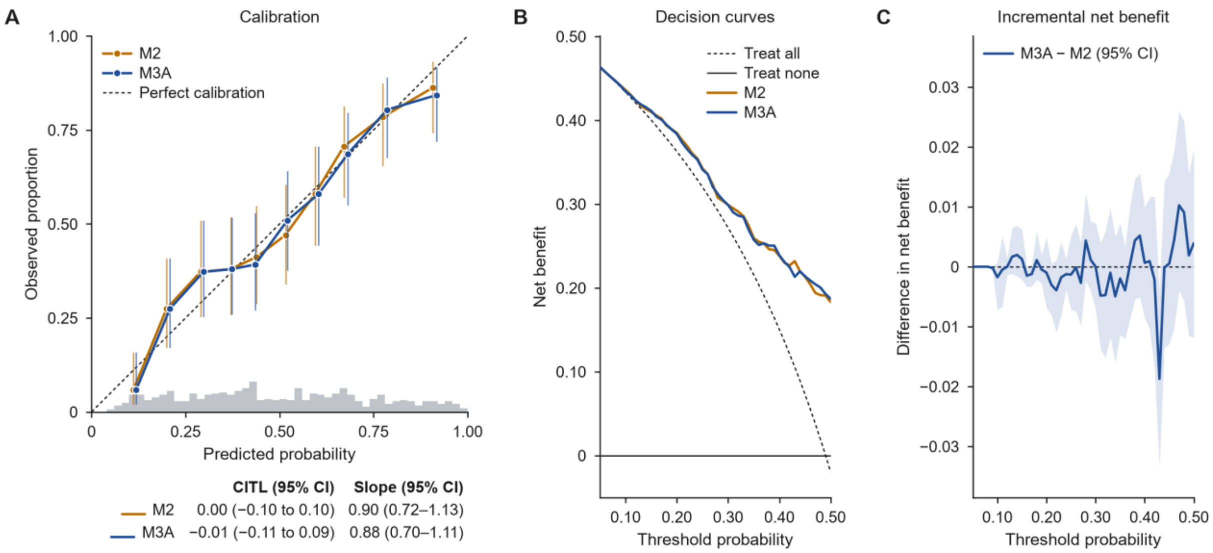


**Supplementary Fig. S1. Calibration and decision curves for the primary comparison in the development cohort.** Out-of-fold predictions of logistic regression M2 and M3A in the development cohort (n=508); panels as in Figure 4.

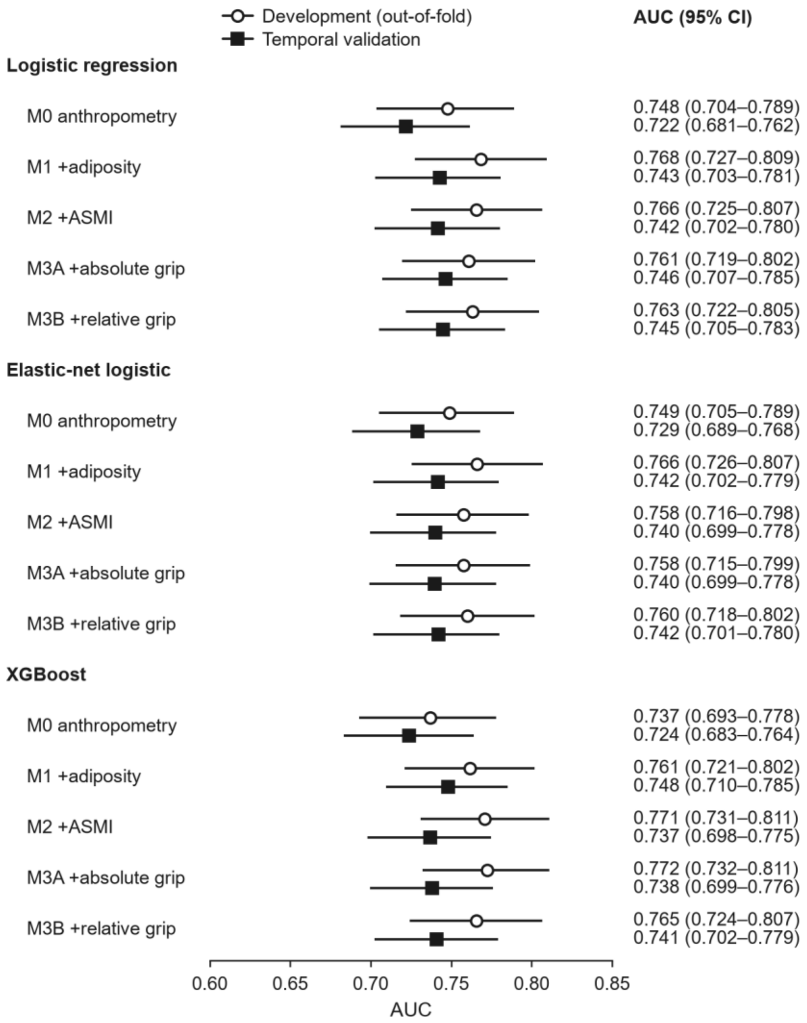


**Supplementary Fig. S2. AUC of all 15 models in both cohorts.** Open circles show development out-of-fold estimates (n=508) and filled squares temporal validation estimates (n=616); lines are 95% CIs from 2000 stratified bootstrap resamples.

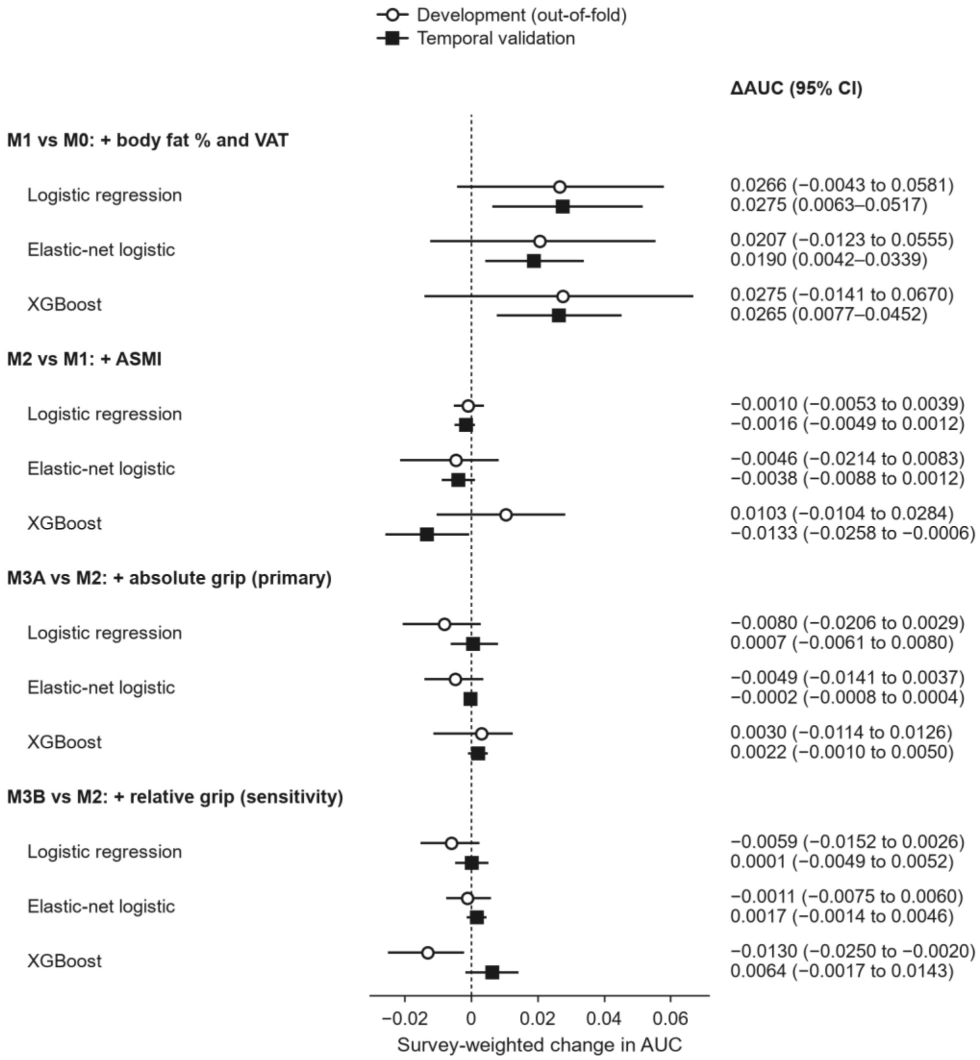


**Supplementary Fig. S3. Survey-weighted change in AUC for the prespecified comparisons.** Weighted with fasting-subsample examination weights; 95% CIs from a Rao–Wu rescaling bootstrap (2000 replicates) that resampled pseudo-PSUs within masked variance strata. Predictions are those of the frozen models; no model was refitted. Rows and symbols as in Figure 3.
